# POST-ACUTE SEQUELAE OF COVID-19: CHARACTERIZATION, COMORBIDITIES, AND BIOMARKERS IN A DIVERSE COHORT

**DOI:** 10.1101/2024.06.20.24308901

**Authors:** Emily L. Struttmann, Anish Shah, Matthew Moreida, Maxwell Rubin, Shanan Immel, Utsav Patel, Bhoomija Chatwani, Shelby Flaherty, Sharon Liu, Marc Theberge, Allison Lockhart, Charlize Nguyen, Jaz Montes, Joshua Katz, Arnaud C. Drouin, Dahlene N. Fusco

## Abstract

**Introduction:** Post-acute sequelae of COVID-19 (PASC) is causing a silent pandemic in the U.S. Gulf South, a part of flyover U.S. where patients are quietly withdrawing from the workforce and largely disconnected from the advocacy resources growing in more affluent regions[1]. To date, there is no clinical test to diagnose PASC and PASC risk factors and etiology remain unclear.

**Methods:** This prospective study investigates PASC alongside pre-COVID-19 medical history, acute COVID-19 course, and a panel of 25 blood biomarkers collected from 100 COVID-19 patients in New Orleans, LA, in a 52.5% Black cohort, providing a unique opportunity to describe PASC symptoms and associations within a comorbidity-rich population. 107 participants recruited from the ClinSeqSer COVID-19 study at University Medical Center (UMC) or Tulane Medical Center (TMC) in New Orleans underwent PASC symptom questionnaires at 3-month intervals. 100 blood samples from patients at their initial post-COVID follow-up visit were analyzed for cardiac, metabolic, inflammatory, coagulation, chemistry, and hematologic markers in a clinical laboratory. Results were analyzed in SPSS for associations with PASC positivity which was defined as presence of three or more new-since-COVID symptoms present at a visit 12 or more weeks after COVID diagnosis. PASC prevalence was also analyzed alongside demographics and past medical history.

**Results:** Enrolled participants ranged from 21-87 years old (median 53, mean 52.1, STD 13.7). 63% of participants were female, 52.5% Black, 44% White, and 3% Asian. 52% of participants were hospitalized during their acute COVID-19 course. Severity of participants’ prior acute COVID was known for most subjects. For 82% of subjects, nasal swab and or saliva SARS CoV-2 qRT-PCR value was known and PCR values did not predict later PASC. Maximum severity scores were assigned to 100 out of 105 participants from whom acute COVID-19 data was collected. On average, patients reported over 5 new-since-COVID symptoms and 75% of patients who completed a questionnaire at time of blood draw were PASC positive. Questionnaire results identified common new-since-COVID symptoms including fatigue (64%), dyspnea (53%), myalgias (48%), trouble concentrating (48%), and memory problems (50%). Over one third of participants reported new-since-COVID arthralgias (34%), headaches (40%), and problems sleeping (40%). For all patients reporting these common symptoms, average frequency and severity of symptoms were reported on a scale of 1 (mild) to 5 (severe) as follows (frequency; severity): fatigue (3.3; 3.3), myalgia (3.4, 3.4), memory problems (3.1, 3.2). Comparison of means analysis indicates that hemoglobin, hematocrit and calcium are lower in PASC positive patients but still within normal range. Analysis of demographics indicates that females in this study are 4.8 times more likely to be classified as PASC positive than males. Serology identified a mild trend toward higher anti-N concentration, and plasma proximity extension proteome detected higher IL-6 and TNF, among PASC vs non-PASC.

**Discussion:** PASC is highly prevalent among post-COVID subjects in this 52.5% Black cohort. A panel of commonly ordered clinical labs was unable to distinguish PASC vs non-PASC subjects, indicating an ongoing need for diagnostic biomarkers relevant across diverse patient groups.

## 1 Introduction

Post-acute sequelae of COVID-19 (PASC), alternatively referred to as Long COVID, has been coined to describe the persistence of SARS CoV-2 infection-related symptoms for months after the acute COVID-19 course. To date, there is no clinical test to diagnose PASC or to predict who is at greatest risk of developing PASC. There are even discrepancies in defining PASC, though the WHO has defined post COVID-19 condition as symptoms lasting at least two months not better explained by an alternate diagnosis [2]. There is still much to be studied regarding risk factors and etiologies of PASC, despite PASC occurring in at least 10% of severe cases of COVID-19 [3]. Many teams, including the collaborative RECOVER cohort in the U.S., are actively researching PASC biomarkers [3–7].

Several clusters of symptoms have been recognized by the U.S. Centers for Disease Control and Prevention (CDC) as being frequently reported among individuals recovering with PASC including changes in mental function, cardiovascular complications, neurological conditions, and persistent respiratory symptoms [8]. These PASC presentations are likely due to neuroinflammation, myocardial inflammation, dysautonomia, and abnormal gas exchange, among other pathologies [3]. Identifying these pathologies has allowed for development of targeted treatment regimens for these common manifestations of PASC, including some improvement of cognition with administration of famotidine or actovegin [9]. Some data suggests that PASC presents as multi-system dysfunction due to stimulation of inflammatory and coagulation pathways as well as ACE2 receptor binding and further immune dysregulation [10]. Mechanistic hypotheses attribute PASC to prolonged SARS CoV-2 shedding in the gastrointestinal tract or blood [7, 11, 12] or mitochondrial dysfunction [13–15]. It is possible that PASC in sub-cohorts with different symptom clusters, or following different virus variant exposures, experience different mechanisms of pathology.

This prospective study aimed to identify correlates of PASC in pre-COVID medical history, acute COVID-19 course, and a panel of 25 blood biomarkers collected from 100 COVID-19 patients in a 53% Black cohort in New Orleans, Louisiana (LA). New Orleans is a region of the US with very high frequency of pre-COVID comorbidities, with LA being ranked among five least healthy US states for multiple health metrics since 1990[16] and regions of LA’s “Cancer Alley” noted to have highest coronavirus death toll per capita in the US at points during the pandemic[17] This study population provides a unique opportunity to describe PASC symptoms and associations within a comorbidity-rich and 53% Black cohort.

## 2 Materials and Methods

### 2.1 Study Design

#### Ethics statement

All subjects provided written informed consent and written HIPAA form prior to enrollment, under IRB 2020-396, Tulane University School of Medicine.

105 participants with past COVID-19, confirmed by clinical nasopharyngeal qRT-PCR test, were recruited from the ClinSeqSer study of acute and post COVID patients receiving care at University Medical Center or Tulane Medical Center in New Orleans, Louisiana. 2 additional participants without history of COVID-19 infection were recruited to serve as negative controls for a total of 107 participants. Clinical and demographic data was extracted from electronic medical records (EMRs) including pre-COVID medical history, acute COVID-19 course, and maximum symptom severity.

At various time points after discharge, participants underwent a PASC symptom questionnaire, physical exam, nasal swab, saliva collection, urine collection, and blood draw at 3-month intervals. Additional blood was collected from 100 patients at their initial post-COVID follow-up visit at Tulane outpatient clinics and analyzed for cardiac, metabolic, and inflammatory markers along with coagulation, chemistry, and hematologic panels. Study design is presented in **Figure 1**. Results of the clinical blood tests were analyzed in SPSS for associations with PASC positivity. For this study, PASC was defined as at least three new-since-COVID symptoms lasting at least 28 days, present at least one month following symptom onset. This initial definition was selected based on the preliminary clinical observation, starting in May 2020, that post-COVID subjects were often complaining of a series of new symptoms (eg, fatigue, impaired sleep, impaired concentration), and is consistent with current CDC definition[18]. For those participants without a known date of symptom onset, date of enrollment in the Clin-Seq-Ser study was used to approximate date of symptom onset as enrollment occurred at the time when a participant sought medical care for acute COVID-19 symptoms. Participants for whom clinical data, PASC survey results, and clinical lab results were all available were included in association analysis (N=99).

**Figure 1.**
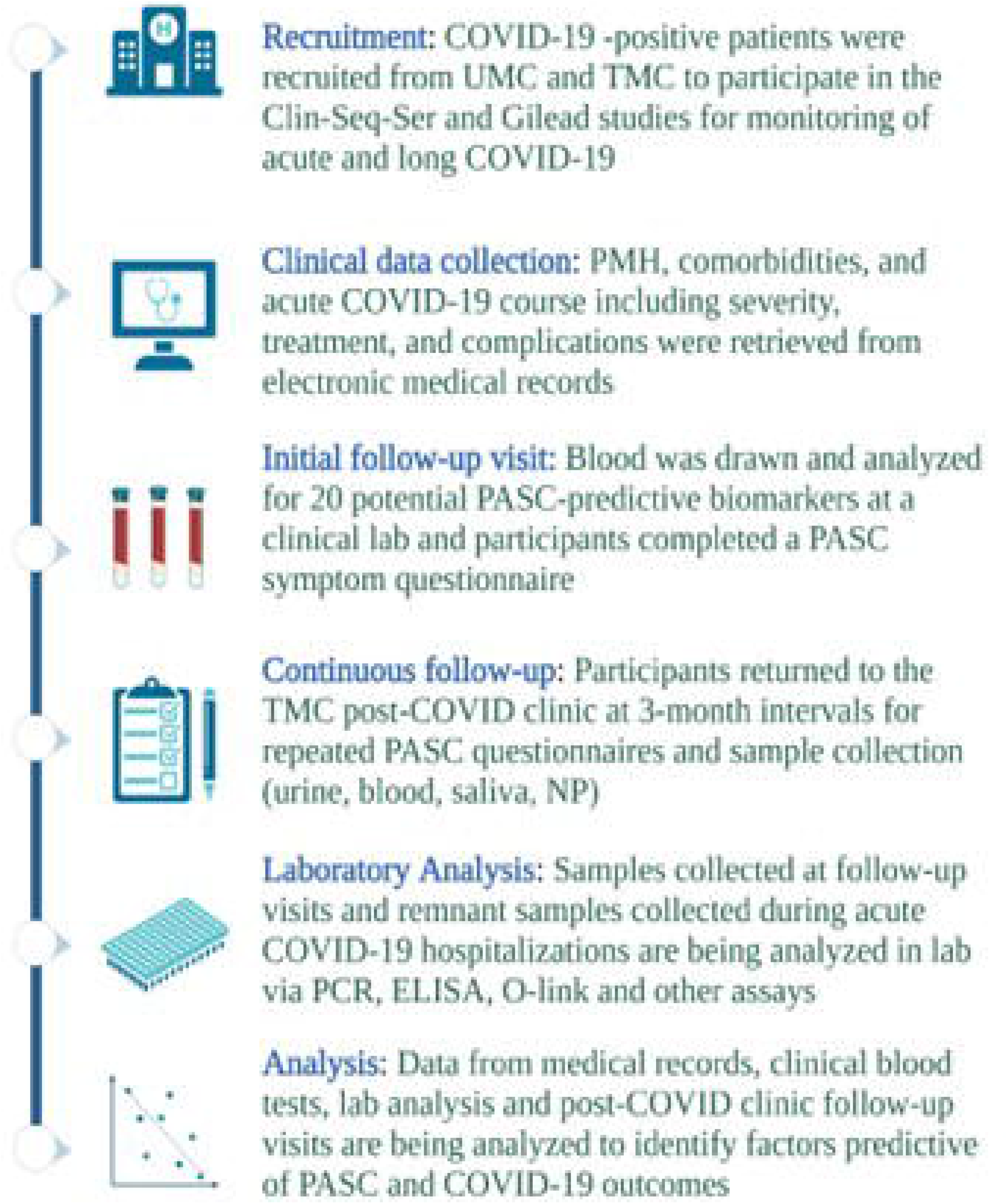

### 2.2 Data Collection and Analysis

A 4-page paper PASC questionnaire was administered to patients at each post-COVID-19 clinic visit to collect PASC symptom data. The questionnaire was collaboratively developed with the CDC and is derived from a previously validated assessment tool for chronic fatigue, endorsed by the CDC. Data was then entered by research personnel from the questionnaire into REDCap for data management and storage. Participants who signed a HIPAA waiver underwent medical chart review of pre-COVID medical history and acute COVID-19 course. Medical record data was entered into a REDCap database which was used for data storage and management. If patients could only recall month of initial positive COVID-19 test or symptom onset, median date of that calendar month was recorded as date of symptom onset and initial positive COVID-19 test. For participants who were asymptomatic during initial infection, data of initial positive COVID-19 diagnostic test was considered to be symptom onset.

#### Statistical Analysis

Demographic descriptive data and odds ratios were calculated using Microsoft Excel. Descriptive and population statistics for laboratory and clinical data were analyzed using IBM SPSS version 29.0.1.0. Variables for which >25% of values were below the limit of detection (LOD) were analyzed as binomial variables (undetected vs. detected) for comparison of means. Variables for which <25% of values were below LOD, but some values were reported as below LOD were analyzed by replacing <LOD value with 0.5(LOD). For descriptive statistics, all values below LOD were analyzed as 0.5LOD.

### 2.3 Laboratory Techniques

#### SARS CoV-2 qRT-PCR

Primers, probes, and positive controls for SARS CoV-2 were acquired from Integrated DNA Technologies, in accordance with the “CDCs Influenza SARS CoV-2 Multiplex Assay (Flu SC2 Assay)”. Positive controls, deionized water, a test negative RNA sample and 8 control samples of known RNA concentration were used on each 384 well plate. These controls served as validation for the specificity of the primers as well as success of the prior RNA extraction. Negative controls allow monitoring for contamination and the control samples allow correlation of cycle threshold (Ct) with known RNA concentration to quantify each sample more specifically. Samples with a Ct less than 38 were considered “positive”.

#### Roche Elecsys Anti-SARS CoV-2 (Roche Diagnostics, Rotkreuz, Switzerland) (Elecsys)

The test was performed on the Cobas analyzer. SARS CoV-2 nucleocapsid (N) antigen was used as a target and detected with an electro-chemilumescence sandwich assay (ECLIA). Results are presented as Cut-off-Index (COI) values and are expressed as qualitative statements (reactive/non-reactive). For result interpretation, a COI<1 is non-reactive and a COI ≥ 1 is reactive. The use of numerical cutoff index values is not specifically recommended by manufacturer however graphs with COI values on Y-axis and sequential samples collected days after reactive PCR on X-axis are presented for several patients in the packet insert. The increase of COI values over time after first PCR positive suggest a relationship between COI values and concentration of anti-N IgG. This presentation method was applied to the current study.

#### Zalgen reSARS anti-SARS CoV-2 IgG nucleocapsid ELISA

Zalgen reSARS anti-N IgG is a semi-quantitative ELISA performed manually. Zalgen assay was performed according to manufacturer’s instruction. All sample and control wells were duplicated, and averages were used for any data calculations. First, average optical density (OD) value for blank wells was subtracted from all other wells, providing “adjusted A450nm” values for each sample. OD values for each experimental well at A620nm were then subtracted from adjusted OD 450nm values, yielding what is referred to as the calculated OD. Controls included a known seropositive sample, a known seronegative sample, a blank, and 5 standards of increasing known concentration. Samples with an OD higher than that of the negative control were considered positive and samples at or below this OD were considered negative. The calculated OD of each standard was then plotted on the Y-axis against their known concentrations in a 4-parameter regression using Gen5. The equation of this generated curve was then used to convert the calculated OD values into IgG concentrations for each sample. Zalgen ELISA data analysis: Lab results with values <LOD were substituted with 0.5 LOD. Per manufacturer’s recommendations, an estimated concentration of anti-N IgG in units/mL was obtained for each sample by plotting the mean OD obtained for each dilution of IgG Reference on the Y-axis against the corresponding IgG Reference (unit/mL) value on the X-axis using curve fitting software [arbitrary OD unit/volume of sample].

#### OLink Plasma and Urine

Seventeen unique plasma and eleven unique urine samples were shipped on dry ice to Olink proteomic where they were analyzed via the Olink® Target 96 panel utilizing the Proximity Extension Assay technology to measure relative concentrations of 92 different inflammatory proteins.

## 3 Results

### 3.1 Cohort Characterization

Enrolled participants ranged from 21-87 years old at time of enrollment (with a median age of 53. The cohort was predominantly female (63%). 52.5% of participants identified as Black, 44% as White, and 3% as Asian/ Pacific Islander (**Table 1**).

**Table 1.**
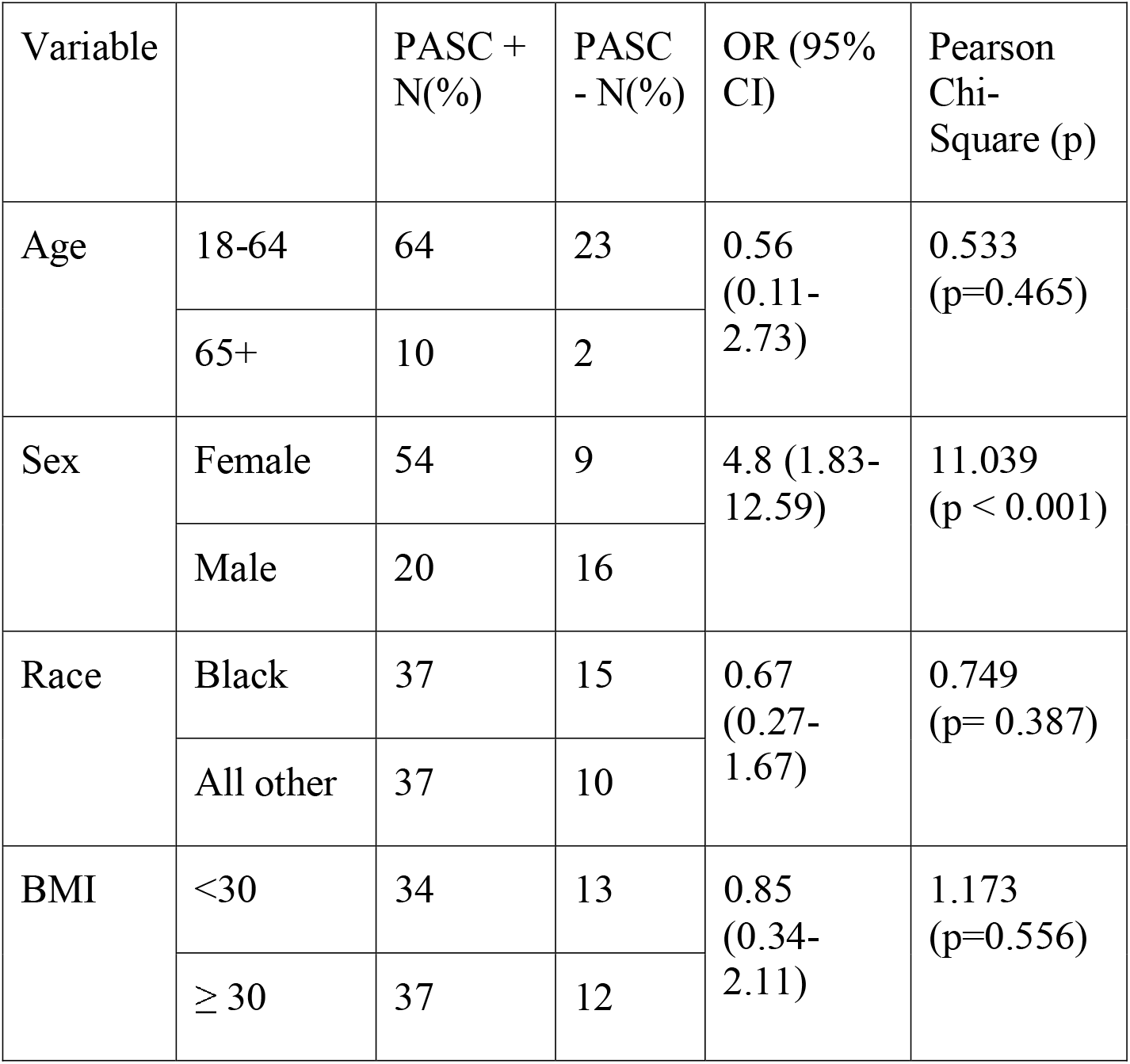
Cohort Characterization. Females in this cohort were 4.8 times as likely to report PASC positivity when compared to males. No significant odds ratios (ORs) were found when age, race, and BMI were stratified by sex

Time between first COVID-19 symptom onset and date of initial post-COVID blood draw was known for all but two participants and ranged from 18 to 663 days with a median of 306 days between symptom onset and clinic visit.. Participants with time between symptom onset and blood draw of less than 28 days were excluded from analysis due to ineligibility to be defined as PASC positive. Participants were seen at post-COVID clinic visits ranging from 1-24 months after symptom onset (**Fig. 2a**). There was great variability in number of study visits and months since symptom onset of these visits between participants, providing insight to PASC symptom prevalence at many timepoints throughout the cohort’s post-COVID course (Fig, 2b).

**Fig. 2.**
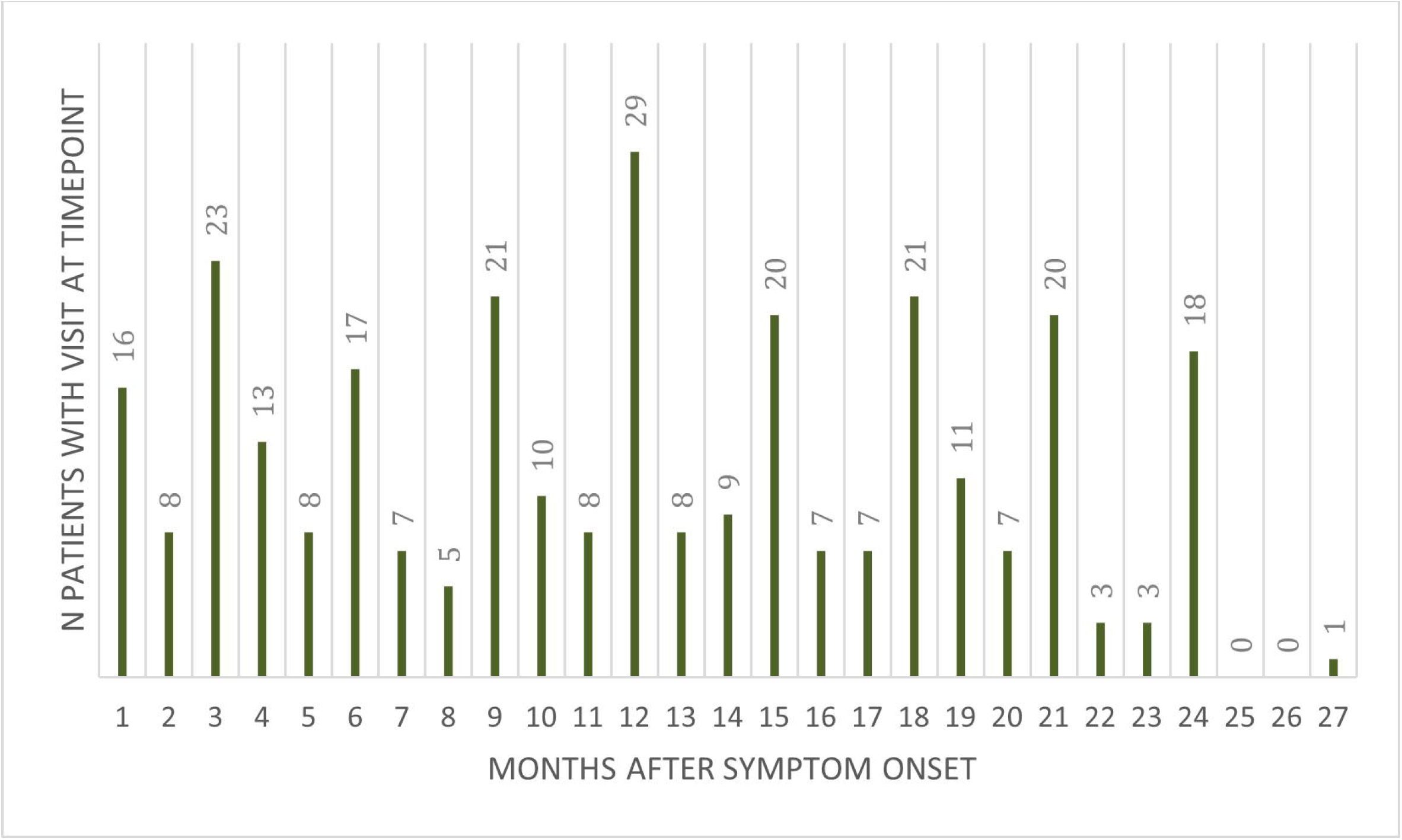

Vaccination status was known for 96 of 107 participants. Of those, 16% were fully vaccinated at the time of initial COVID-19 infection. Patients were considered fully vaccinated if they received 1 dose of Johnson and Johnson or 2 doses of Pfizer or Moderna vaccines at least 14 days prior to acute COVID-19 symptom onset. 40 out of 74 participants (54%) who reported their response to COVID-19 vaccination experienced appreciable side effect, defined as new onset symptoms other than arm pain following vaccine administration.

Past medical history was known for 106 of 107 participants. 93.4% (N=99) had a documented pre-existing medical condition. Past medical history of the cohort was enriched for the following medical conditions: hypertension (31.1%, N=33), other cardiovascular conditions (22.6%, N=24), pulmonary conditions (26.4%, N=28) (**Fig.3**). There were no significant correlations between past medical history and prevalence of PASC in this cohort (**Table 2**).

**Fig.3.**
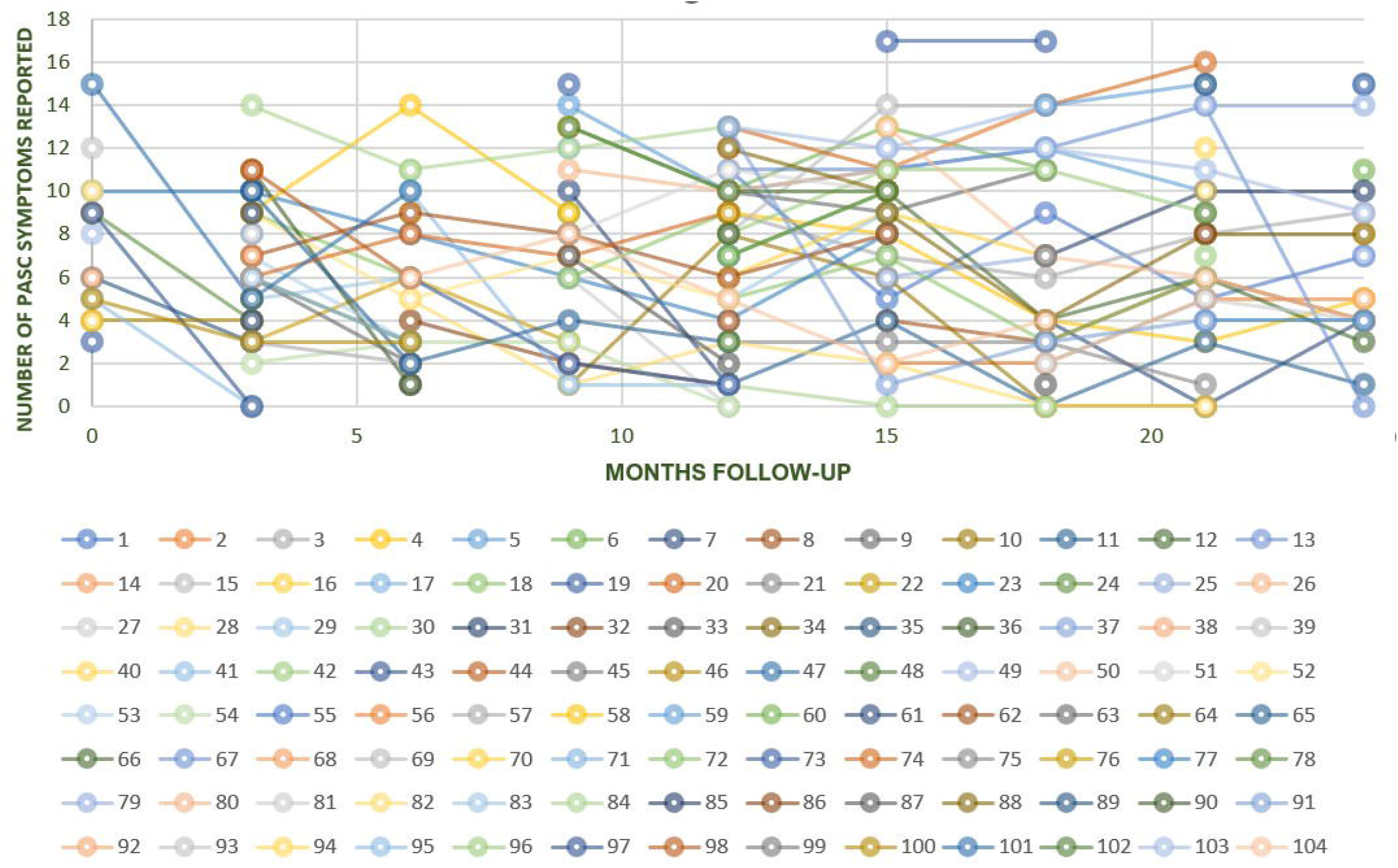

**Table 2.**
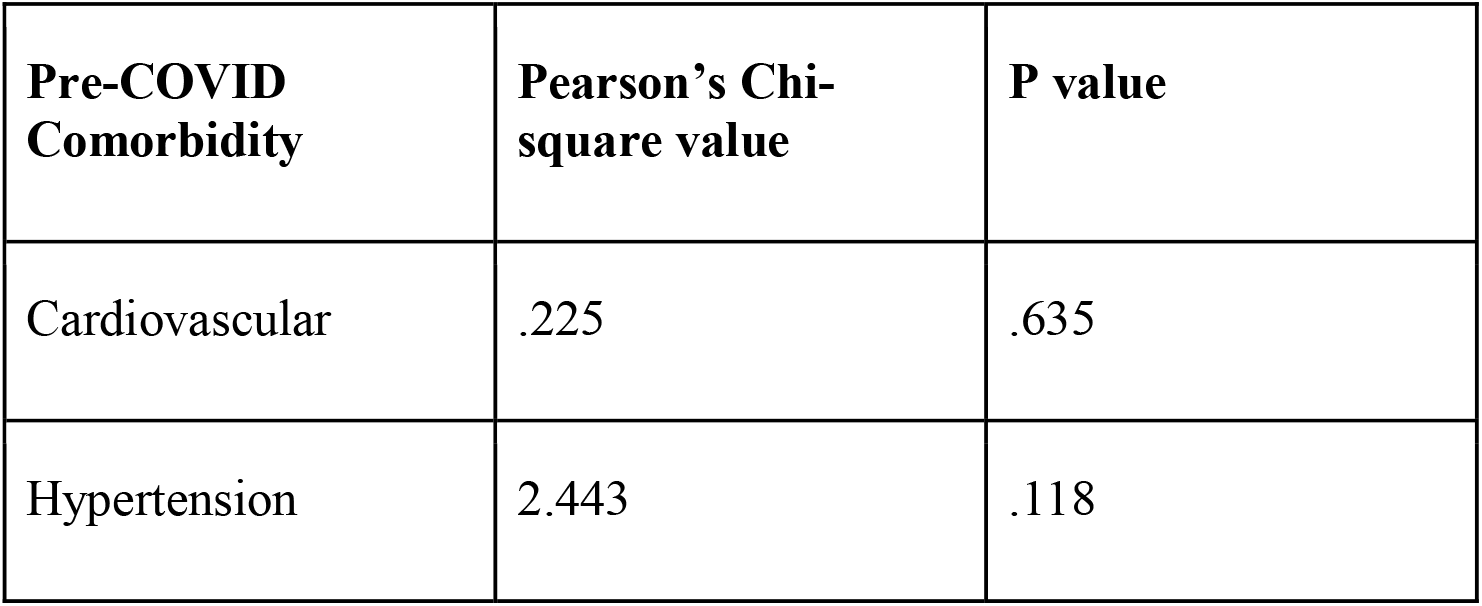
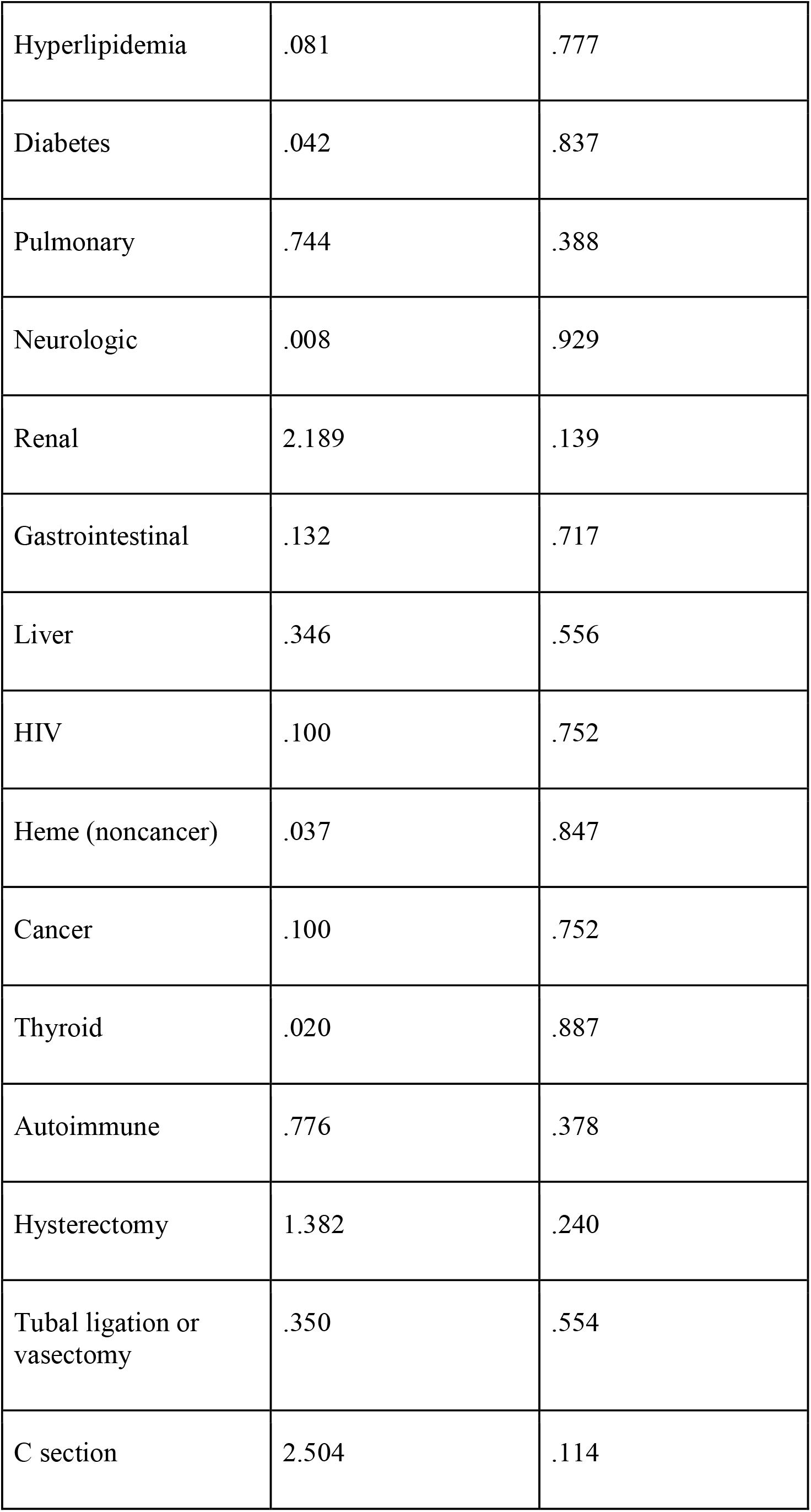
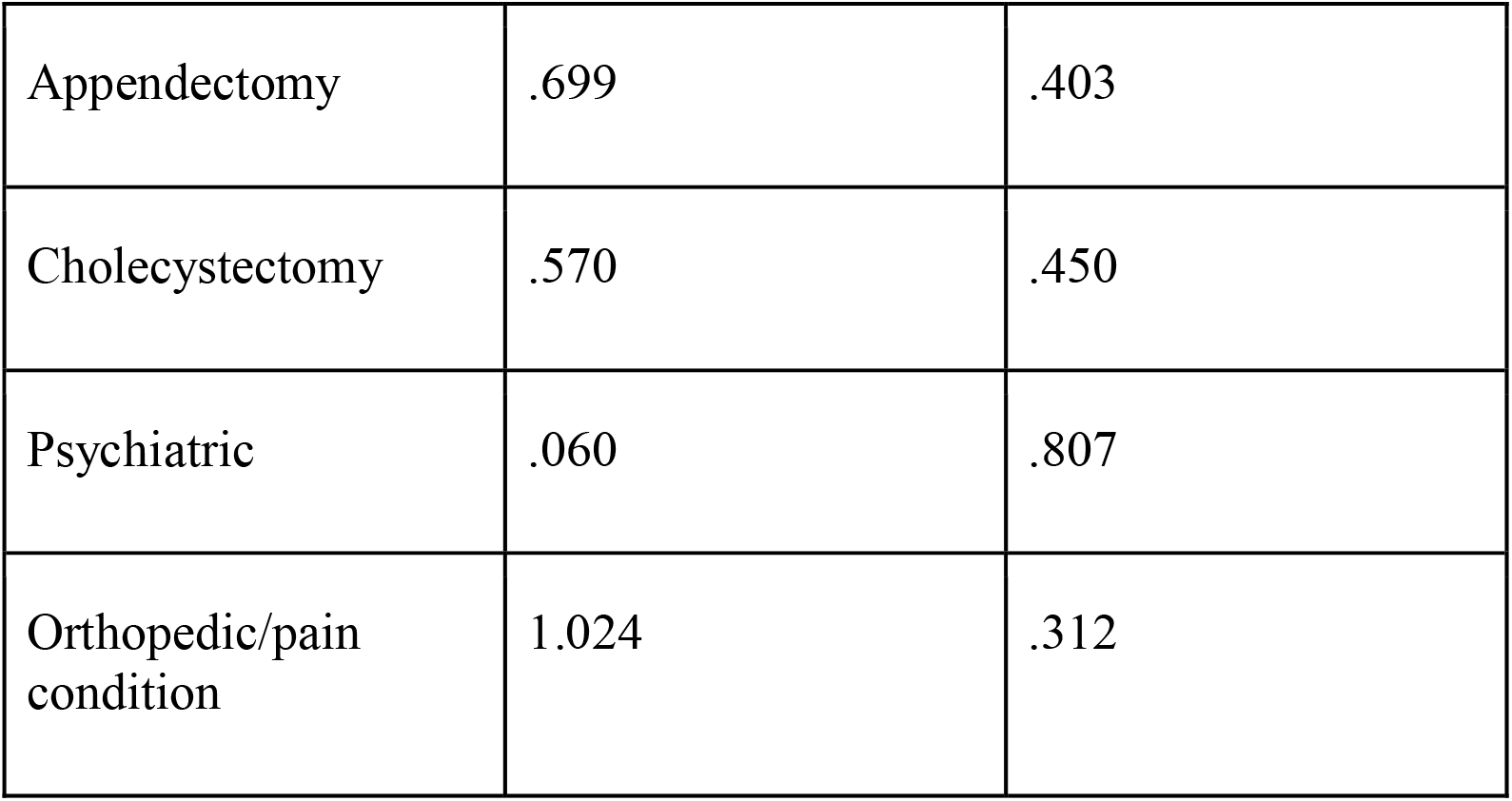
Chi square analysis of comorbidities/past medical history (PMH) and positivity for post acute sequelae of COVID-19 (PASC)

Severity scores were assigned to 102 participants for whom records of acute COVID-19 course were accessed (**Fig. 4**). Severity scores were based on the intensity of respiratory interventions required in management of acute COVID-19 infections (**Table 3**). 47% of participants were treated for acute COVID-19 in the outpatient setting exclusively. The average severity score for this cohort was 5.3, corresponding to an inpatient hospital stay without the use of supplemental oxygen. The average severity of acute COVID-19 course was significantly higher among PASC negative participants (**Tables 4a and 4b**).

**Fig. 4.**
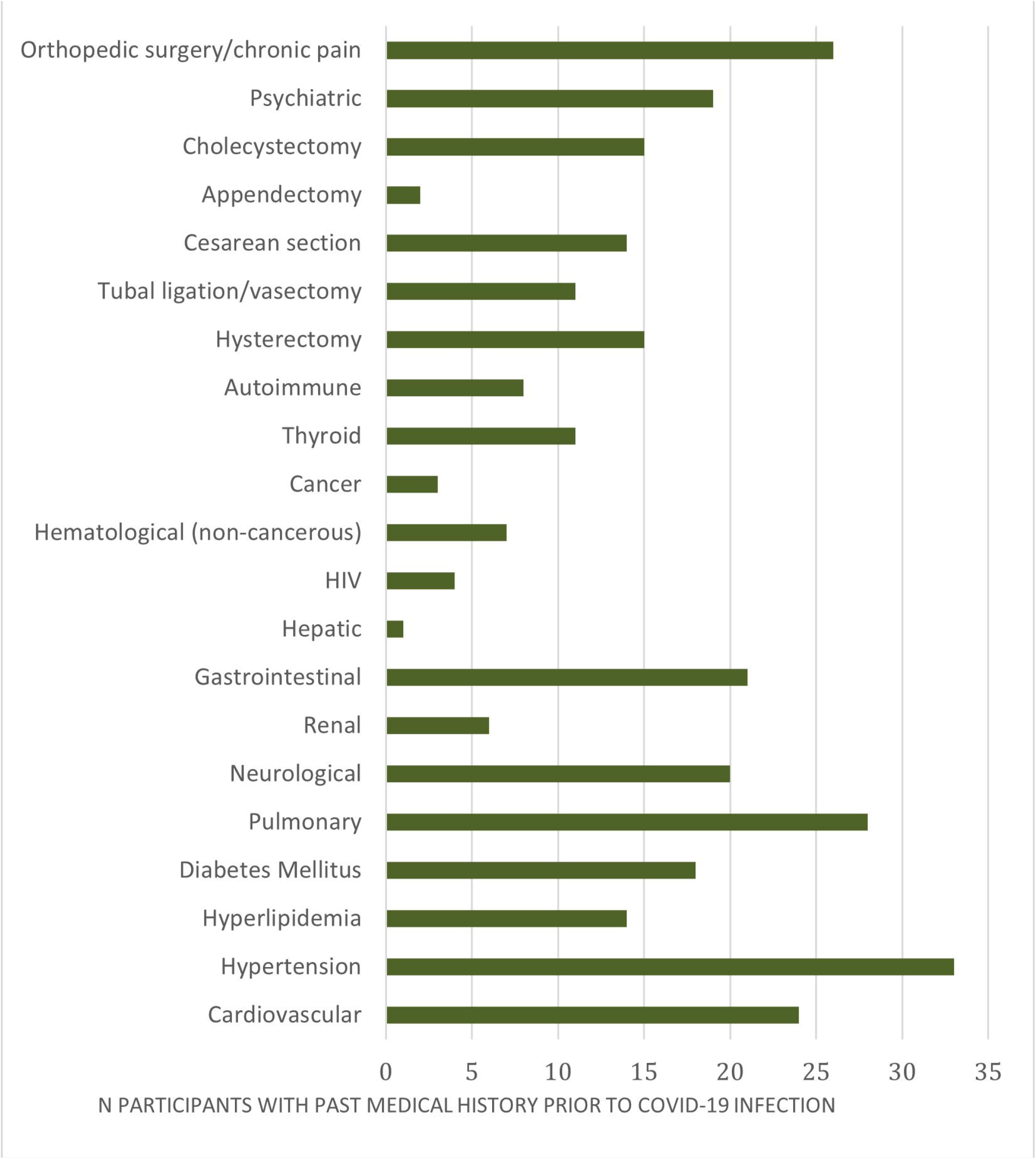

**Table 3.**
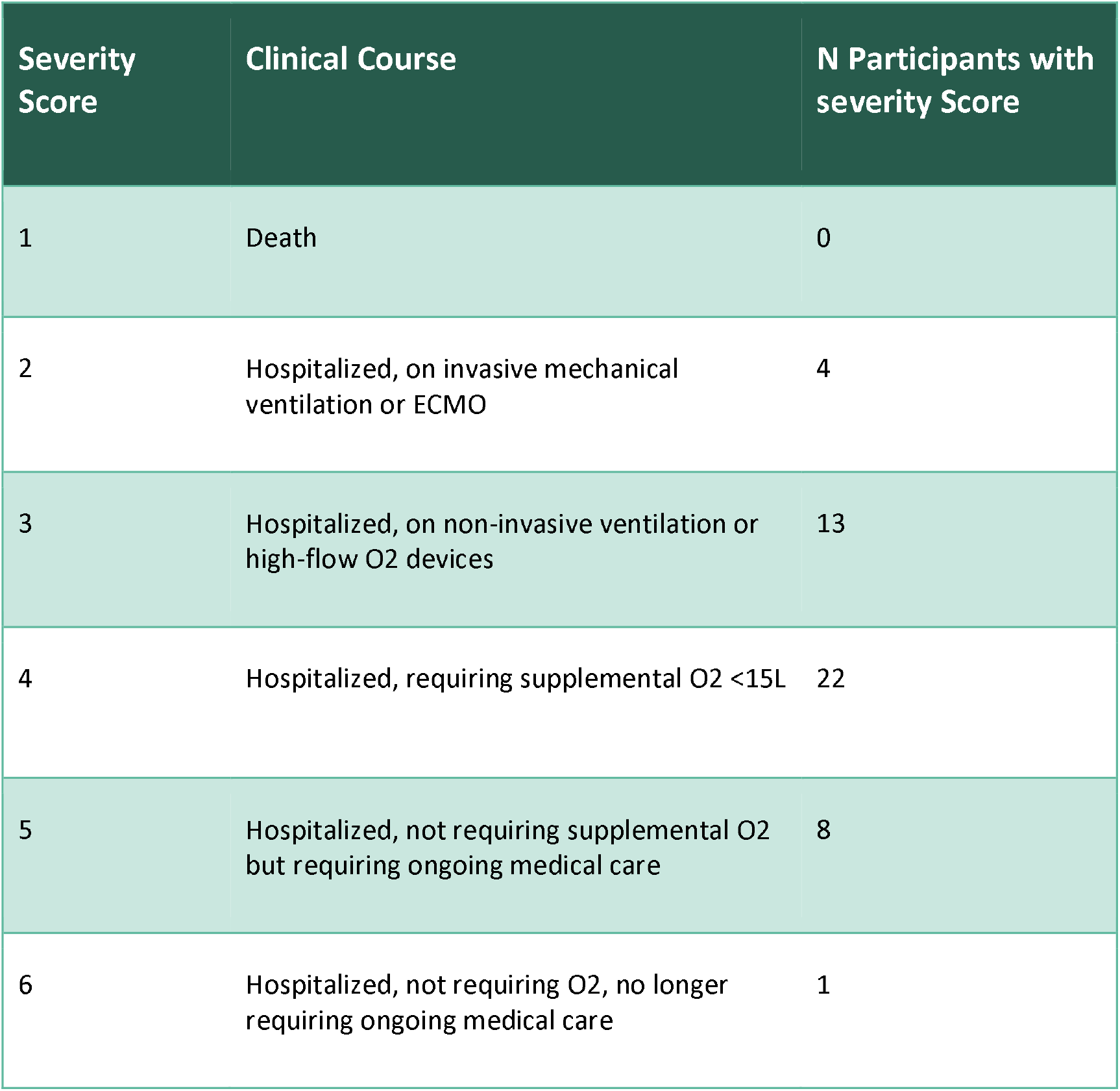
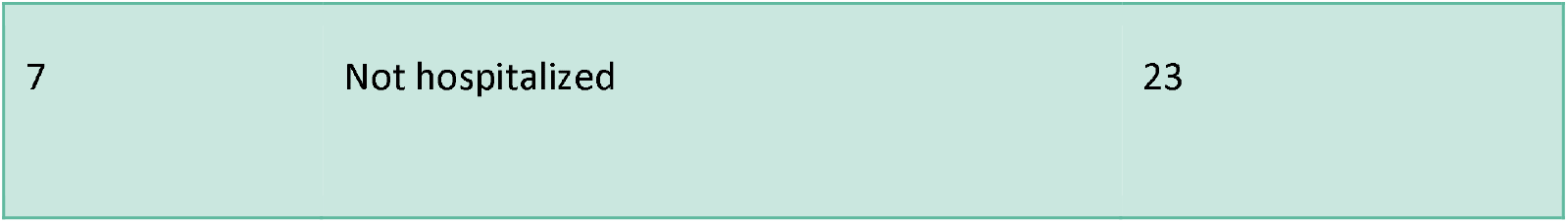
Severity of acute COVID-19 of post-COVID study subjects.

**Table 4.**
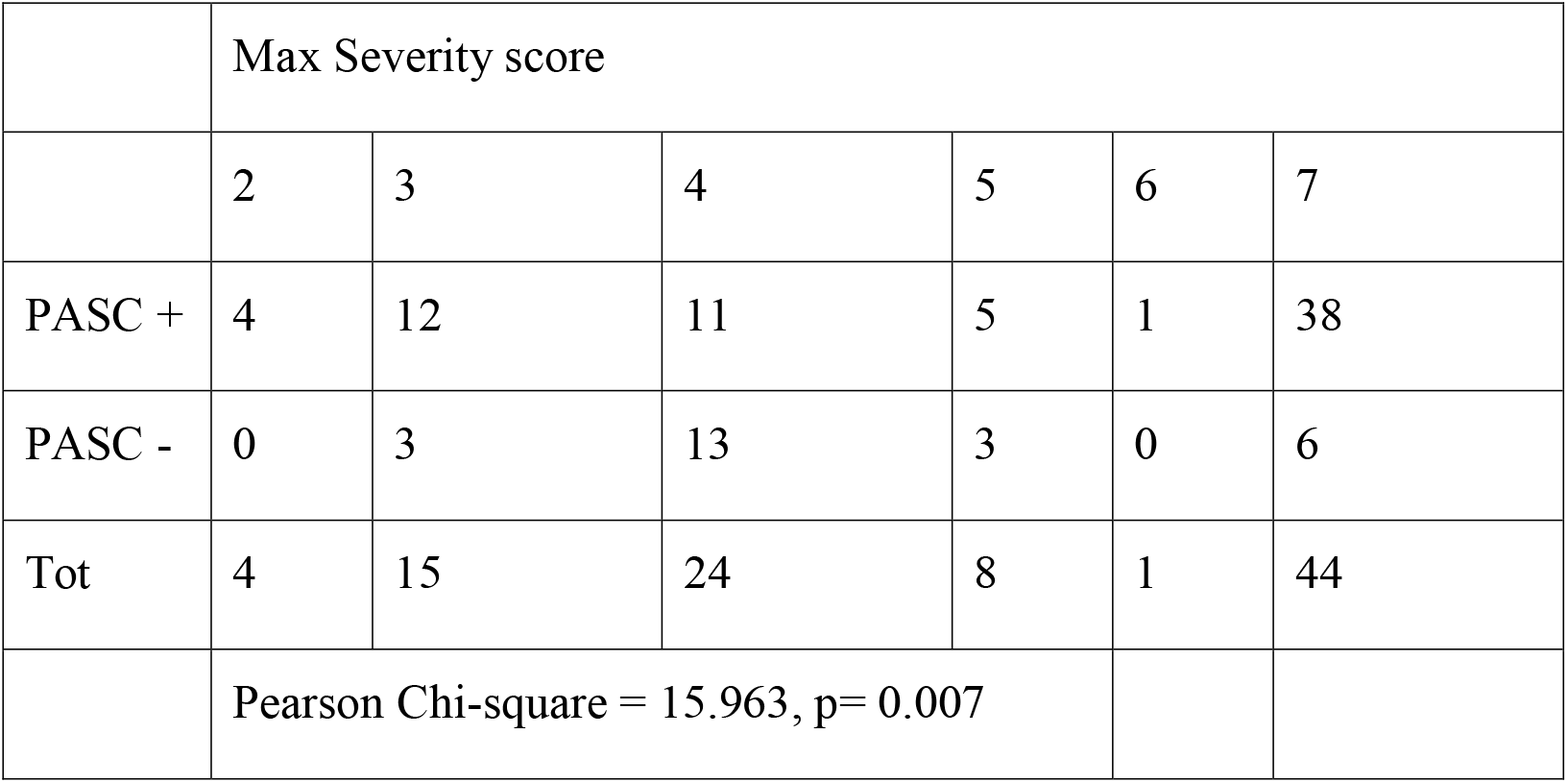
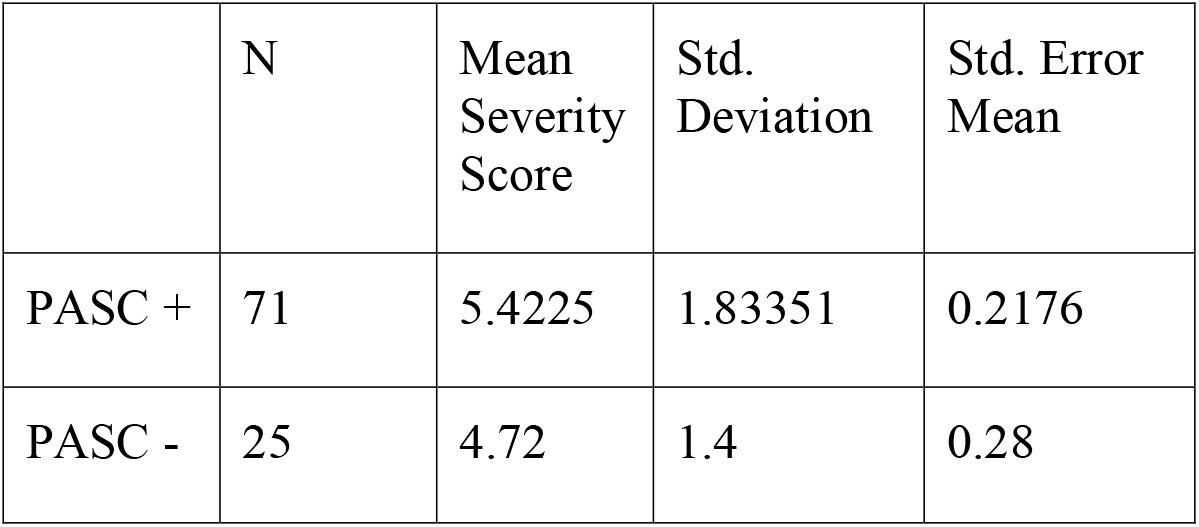
a: Pearson’s Chi-square analysis of Maximum Severity Scores, b: Independent samples T-test of Maximum Severity Scores

### 3.2 PASC Presentation in Post-COVID Clinic Visits

On average, post-COVID study subjects reported over 5 new-since-COVID symptoms with varying frequency and severity. 75% of subjects who completed a questionnaire at the time of blood draw were considered PASC positive.

#### PASC Symptom Prevalence

Results from PASC questionnaire identified common new-since-COVID symptoms reported by participants including fatigue (64%), dyspnea (53%), myalgias (46%), trouble concentrating (48%), and memory problems (50%) (**Fig.5**). The majority of subjects in this cohort presented during pre-Delta and Delta predominant SARS CoV-2 variant circulation, with roughly equal percent PASC for each variant (74% among pre-Delta, 72% among Delta). Throughout the course of the study subjects communicated that study questionnaires were not capturing all of their new-since-COVID symptoms, citing that additional symptoms that should be included in future questionnaires included tinnitus, hair loss, chest pain, food allergies including anaphylactic allergies, and marked alcohol intolerance.

**Fig.5.**
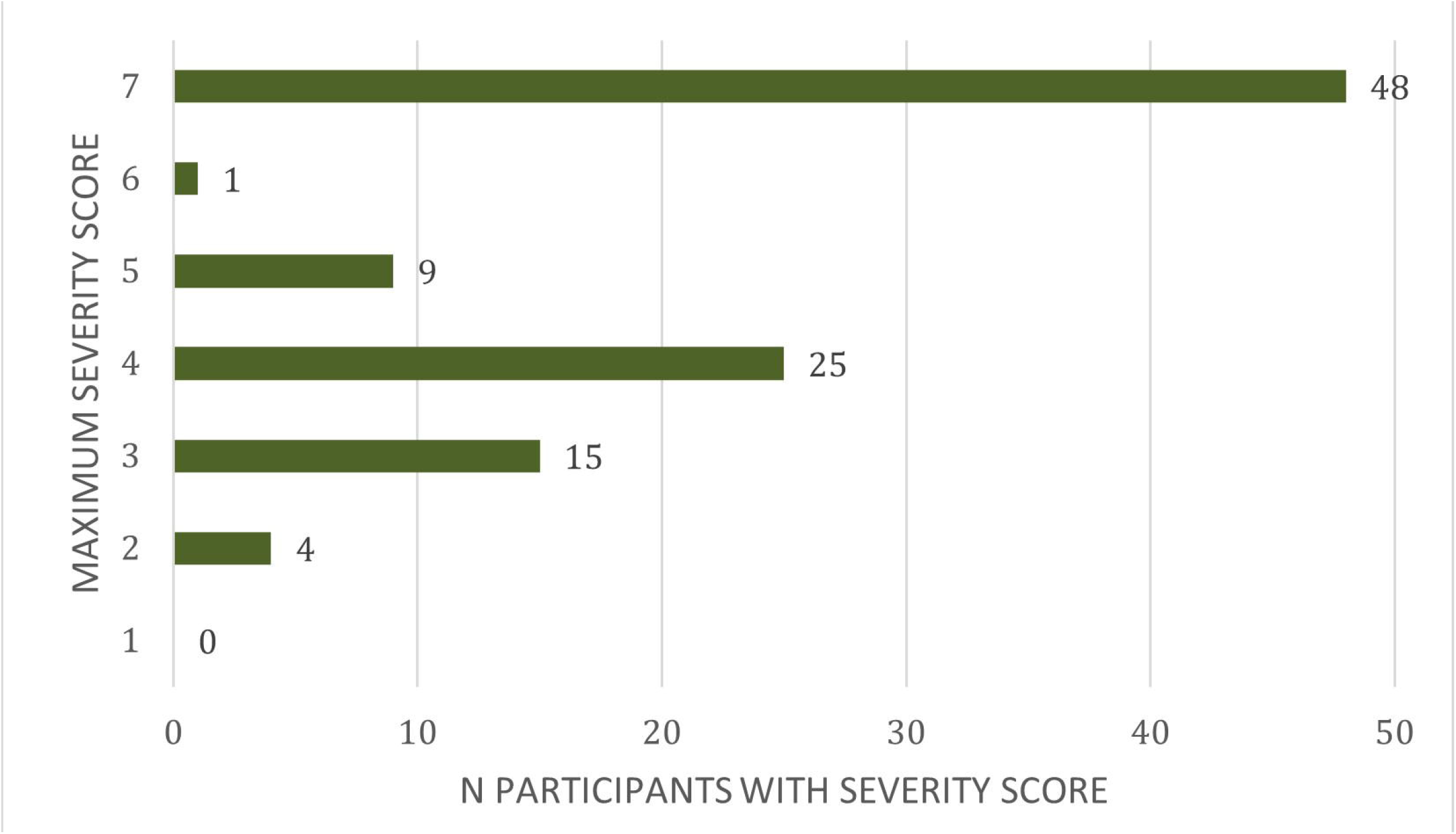

#### PASC Symptom Frequency

On average, participants reported experiencing the above PASC symptoms between “some of the time” and “most of the time.” The symptom reported with the lowest frequency was dizziness/fainting (2.4/5) and the symptom reported with the highest frequency was sleep problems (3.6/5) (**Fig. 6**).

**Fig. 6.**
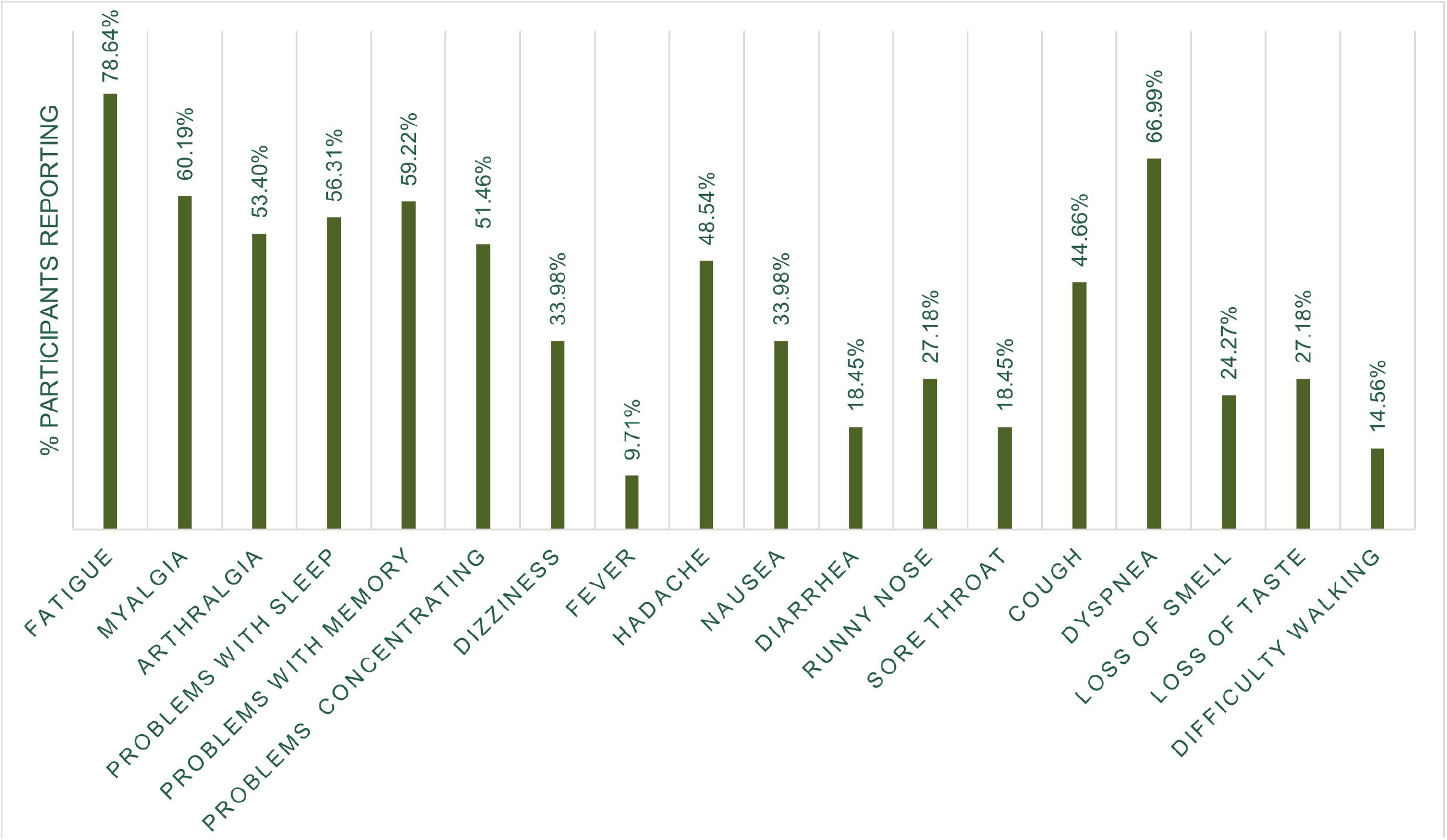

#### PASC Symptom Severity

Average self-reported PASC symptom severities ranged between moderate and severe. The lowest reported severity was for dizziness/fainting (2.9/5 average severity) and the highest reported severity was for problems with sleeping (3.5/5 average severity) (**Fig. 7**).

**Fug. 7.**
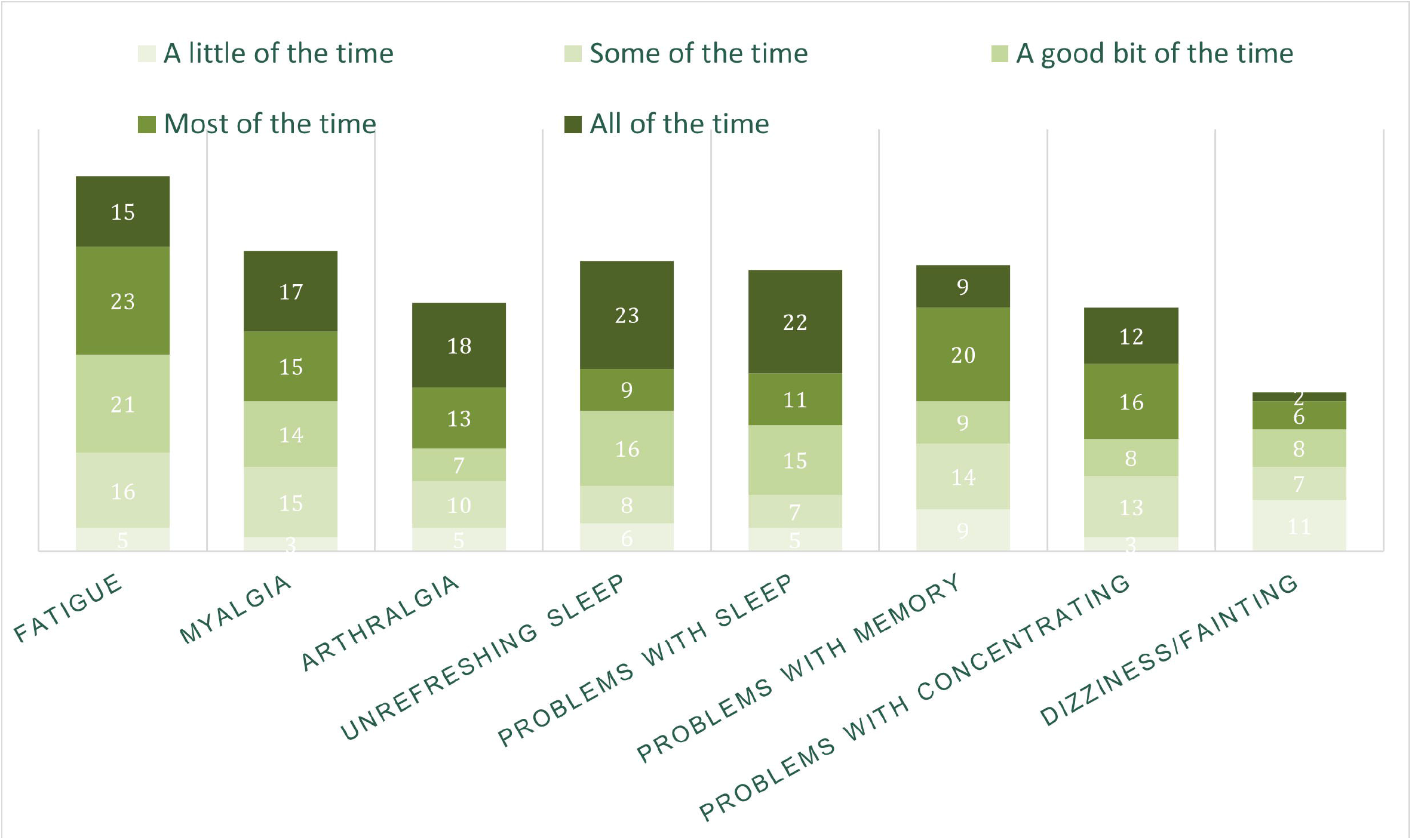

#### PASC Symptom Persistence

The majority of presenting PASC symptoms persisted at more than one clinic visit. Of participants who had at least two clinic visits and reported fatigue at their first visit, 87% reported fatigue at a subsequent visit. Similarly, 83% of participants with multiple clinic visits who reported issues with concentration at their initial clinic visit reported concentration issues during at least one subsequent clinic visit at 3 month timepoints. New-since-COVID fatigue and concentration issues were reported by a substantial proportion of the PASC cohort at every timepoint up to two years after symptom onset (**Fig. 8**).

**Fig. 8.**
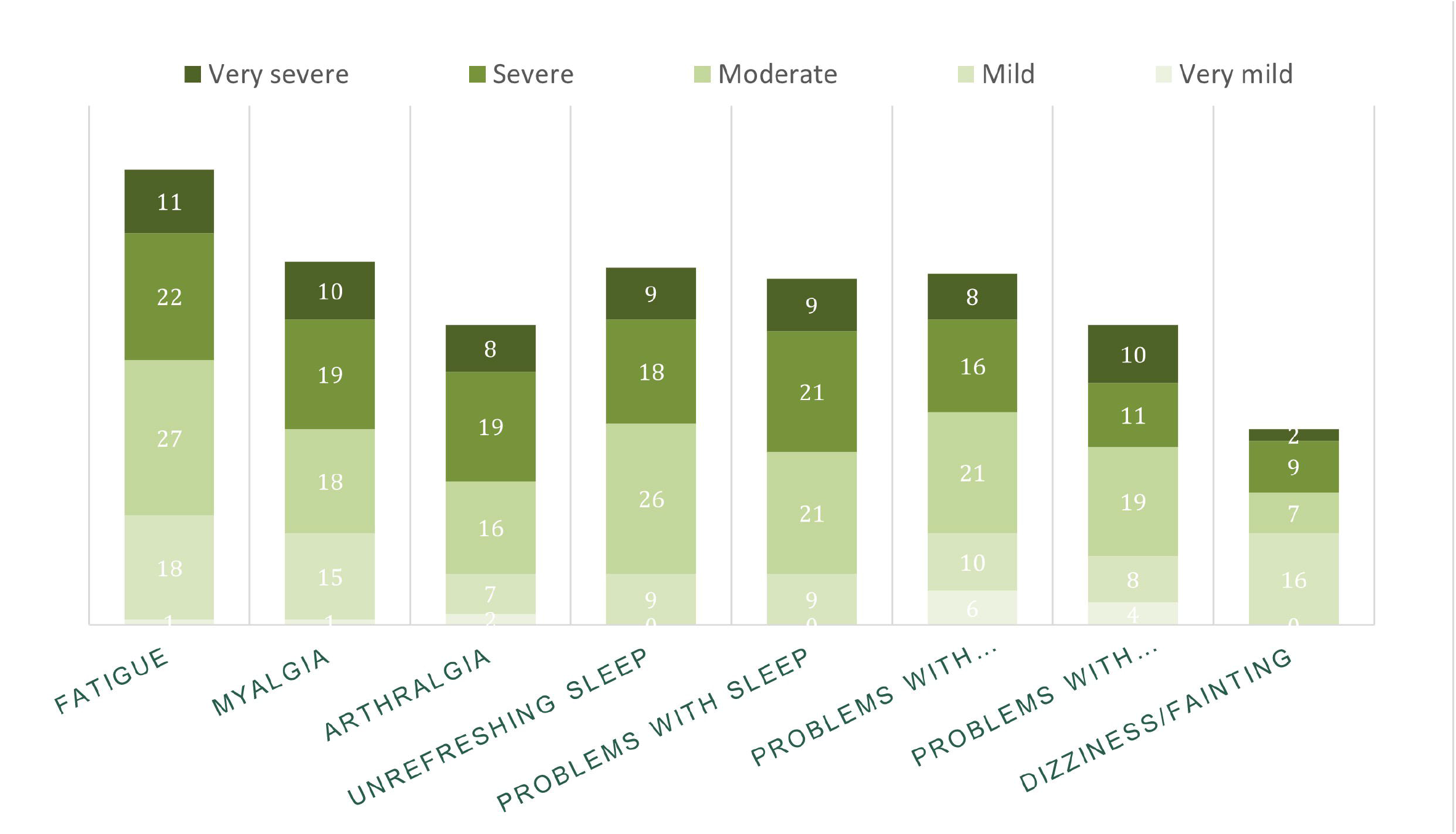

**Fig. 9.**
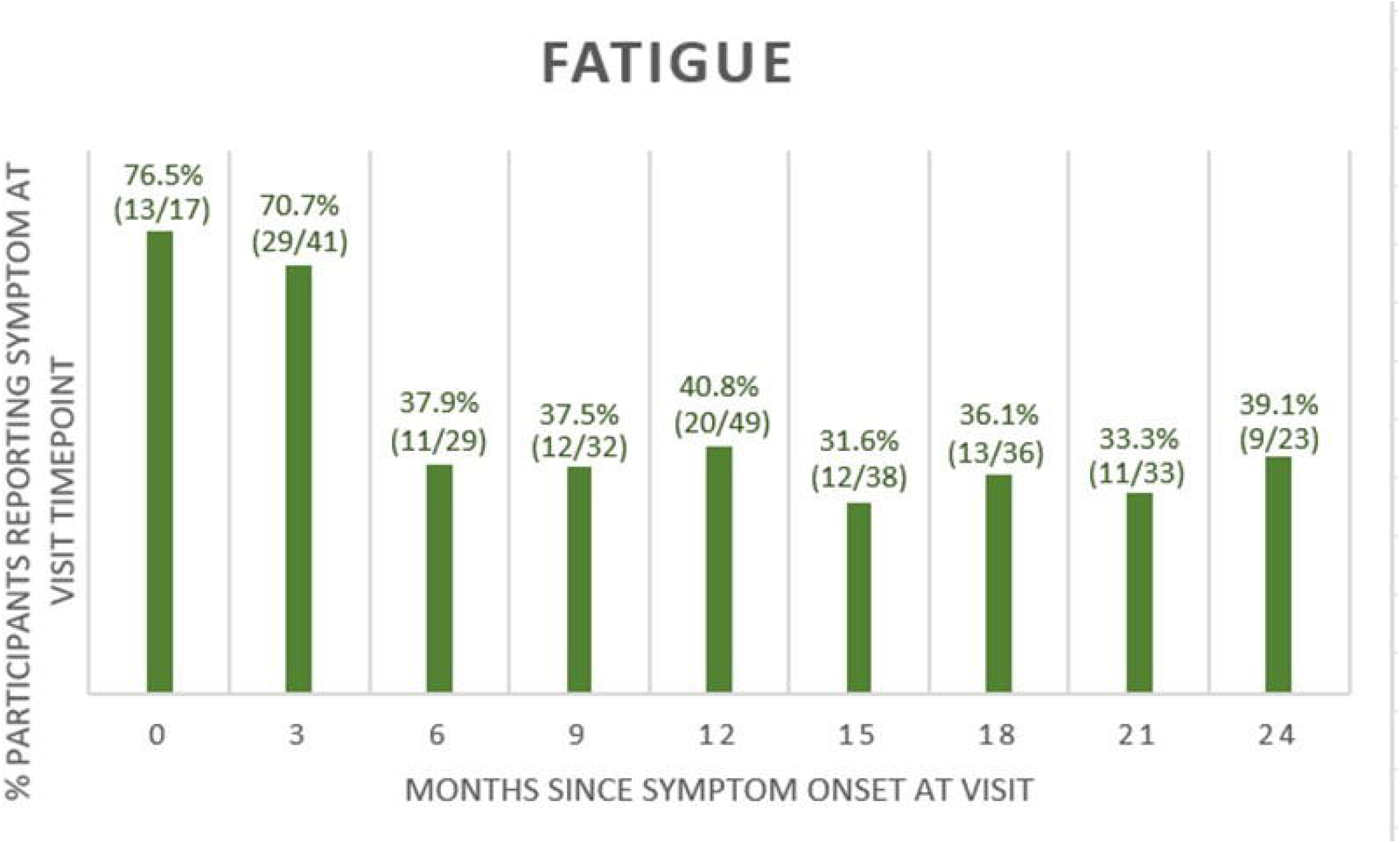

**Fig. 10.**
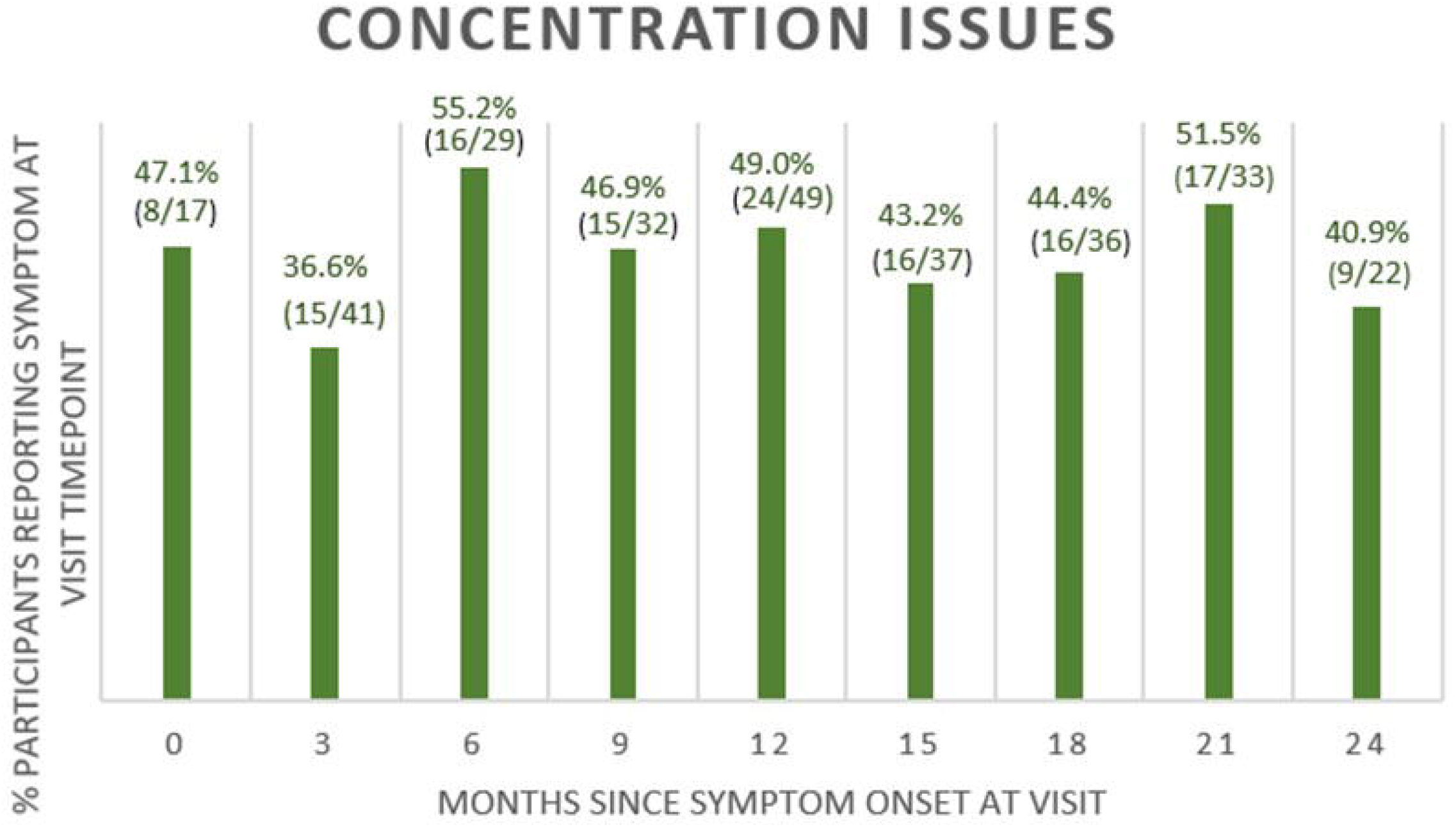

### 3.3 Clinical Laboratory Results

Comparison between PASC positive and PASC negative subjects revealed that hemoglobin, hematocrit, and serum calcium levels are significantly lower among PASC positive subjects, but still all within normal range (**Table 5**). Likewise, detectable C-reactive protein levels were not found to be correlated with PASC positivity (X^2=^ 0.547, p=0.460). Low-sensitivity troponin levels were obtained for 65 participants. All but three participants had low-sensitivity troponin levels <0.015. One PASC-negative and two PASC positive participants had troponin levels detectable above 0.015.

**Table 5.**
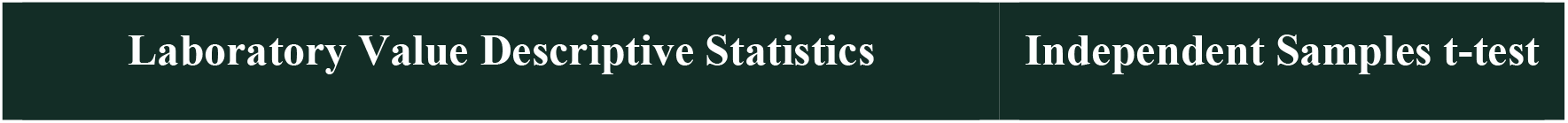
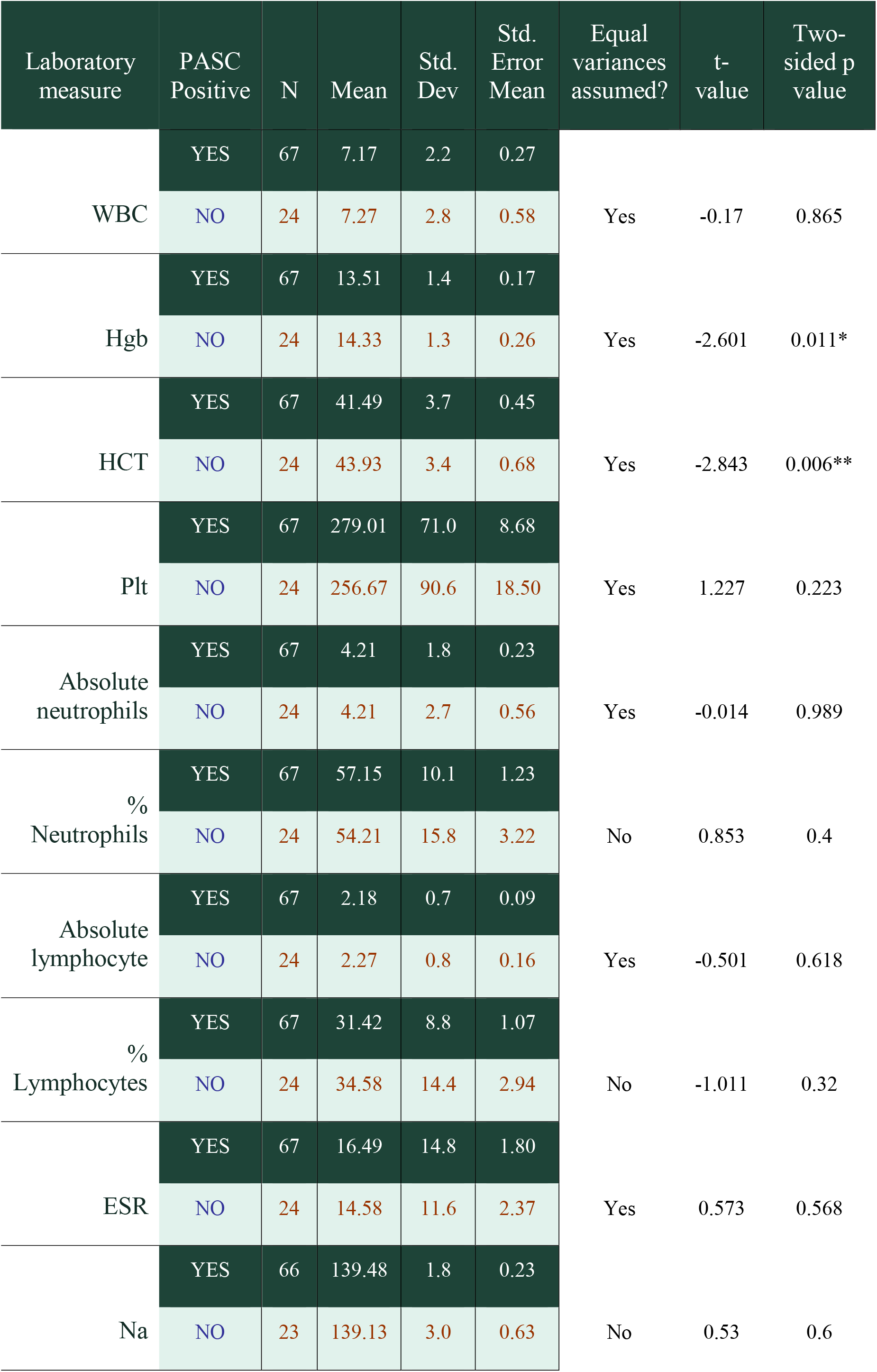
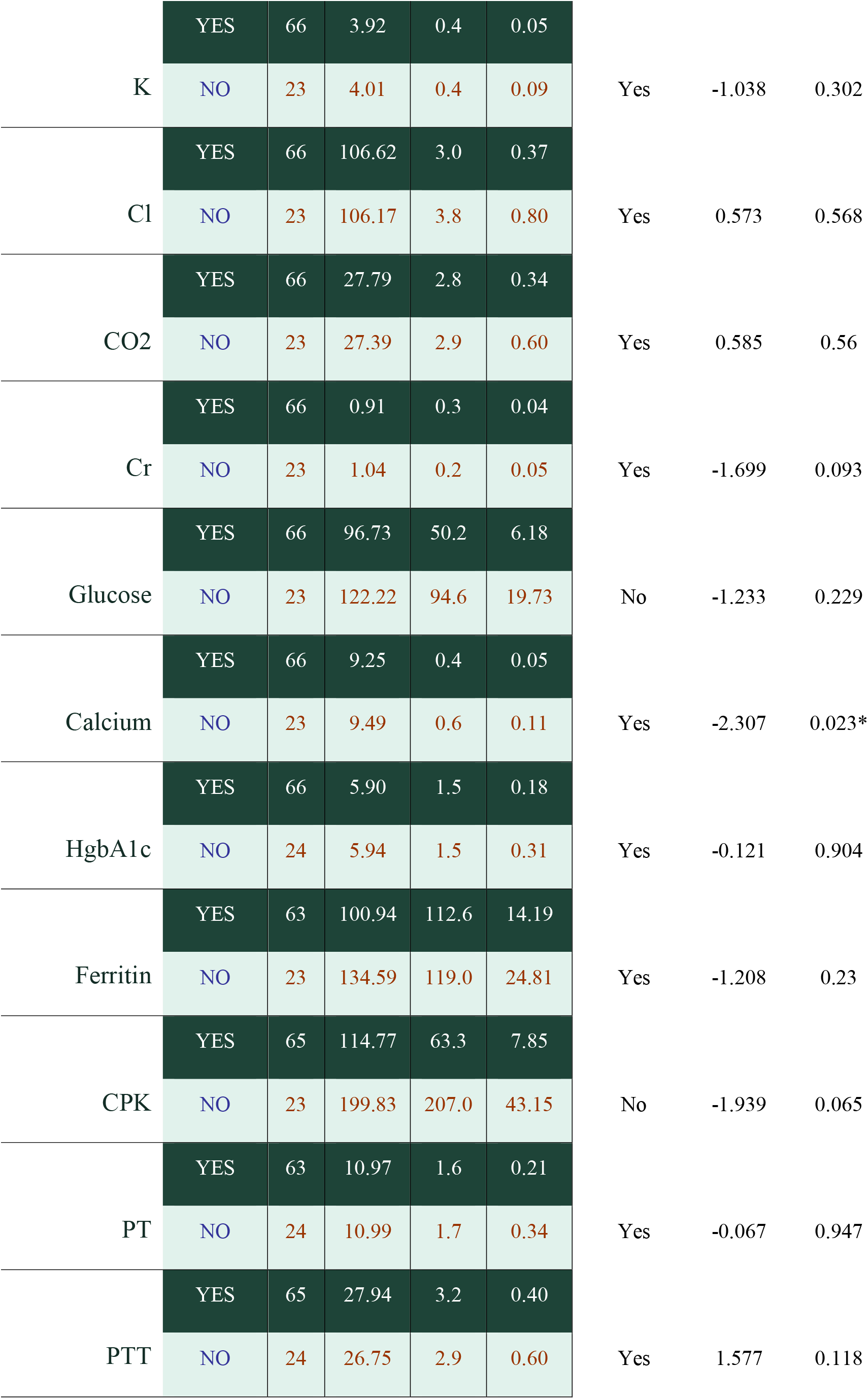
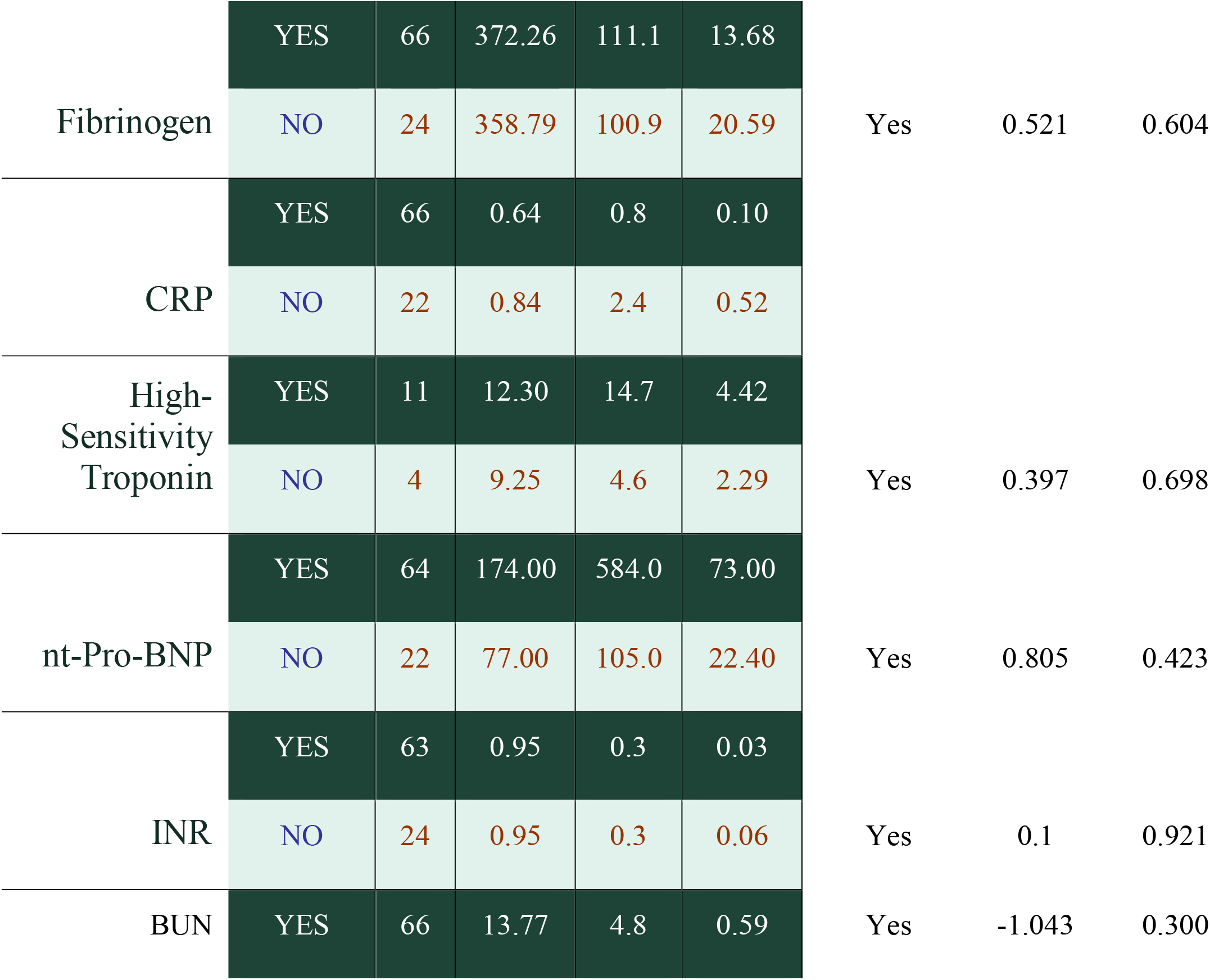
Laboratory Value Descriptive Statistics.

### 3.4 Research Laboratory Results

For a subset of subjects, initial ClinSeqSer enrollment occurred during acute COVID, and included SARS CoV-2 qRT-PCR of nasal and or saliva samples during acute infection as well as possible post-COVID nasal/saliva samples. Other subjects had post-COVID respiratory samples only. Sample collection was optional at all visits. In addition, a subset of subjects had post-COVID SARS CoV-2 serology performed.

#### Respiratory SARS CoV-2 qRT PCR results

SARS CoV-2 qRT-PCR results for nasal (**Fig. S1**) and saliva (**Fig. S2**) samples are presented for individual subjects vs days from symptom onset to sample collection. If a Ct value of 32 is used as the cutoff for negative values, no significant nasal swab or saliva shedding of SARS CoV-2 was detected in this cohort beyond 28 days from symptom onset. Acute COVID-19 PCR cycle threshold (Ct) values were similar for PASC positive and PASC negative participants (N1 p=0.36, N2 p=0.396). Results of acute COVID qRT PCR tests (≤ 28 days) were compared between subjects who were later PASC vs non PASC. This comparison identified no significant difference between PASC vs non-PASC nasal or saliva PCR results for subjects 0-7 or 8-28 days from symptom onset **(Fig.** S3).

For 13 of the post-COVID subjects, a respiratory sample (nasal or saliva) had been obtained during acute COVID-19 and fully sequenced, with results posted on GISAID **(Table S1**). In order to approximate which variant of concern (VOC) caused acute infection in the larger cohort, CDC data was used. Initial COVID infection for the cohort, based on CDC data for general variant circulation time periods, included 64 pre-Delta, 34 presumed Delta, 5 from a period of Delta/Omicron co-circulation and 2 presumed Omicron (**Fig. S4**). Percent PASC positive was: pre-Delta: 74% (46/62), Delta: 72% (23/32), Delta/Omicron cocirculation: 5/5 (100%) (**Fig. S4**). The 2 Omicron subjects did not have questionnaire response data. VOC estimates from CDC data matched VOC for all thirteen subjects who had actual sequencing data.

SARS CoV-2 strains responsible for infections in this cohort mostly spanned from pre-Greek through Delta. Viral genome sequencing results were obtained for 13 subjects (**Table S1**).

#### Serum anti-SARS CoV-2 nucleocapsid ELISA data (Roche and Zalgen)

For Roche/Zalgen serum anti-N data, data from dates other than visit of clinical lab draw were included. PASC/ non-PASC groups were compared for age, severity of initial acute COVID, time between onset and collection, presence of anti-N IgG results, average of COI and arbitrary unit of OD/mL and kinetics of anti-N IgG presence / intensity over time since onset. 6/72 PASC (8%) and 3/25 non-PASC (12%) patients had no sample collected / no Ab results available.

#### Roche anti-SARS CoV-2 nucleocapsid data

Groups of non-PASC and PASC patients had respective proportions of qualitative positive anti-N IgG of 96% vs 85%, and no significant difference of average age (54 vs 52), initial acute COVID severity score (5 vs. 5), or time elapsed between symptom onset and sample collection (292 vs. 334 days).

#### Zalgen anti-SARS CoV-2 nucleocapsid data

Using the Zalgen test, the proportions of anti-N IgG positive results were 97.9% and 77% in non-PASC and PASC groups respectively, with the same trend of higher percent positive anti-N among non-PASC as seen with the Roche anti-N.

#### Zalgen vs Roche anti-SARS CoV-2 nucleocapsid data

Each sample was tested in parallel by Elecsys^R^ Roche / ECLIA technique and by the Zalgen re-SARS anti-N ELISA. 8 samples displayed discrepant results, including 6 positives with Roche/negative with Zalgen. In those 6 the average COI value was low (12) vs. an average COI value of 82 in samples found positive by both techniques. It is tempting to assume that COI values and IgG levels reported by semi-quantitative ELISAs are comparable. We found however no correlation between COI values (Roche) and OD/mL (Zalgen) (r^2^= 0.55), as reported by others [19] confirming that each test result can likely only be compared within the same platform. For subjects with qualitative positive COI, we further compared COI values of PASC vs. non-PASC patients within time periods after onset. Interestingly, COI values showed a trend toward higher values in PASC vs. non-PASC patients between 61-120 days (141 vs 94) and lower after day 120 (64 vs. 84) **(Fig. S5).**

#### O-link Urine and Plasma cytokine levels were obtained for seventeen participants

Though interpretation of these results is limited as many values are below LOD for this assay, IL-6 and TNF values were significantly higher for PASC positive participants (**Table S2**).

## 4 Discussion

PASC is highly prevalent among post-COVID subjects in this 52.5% Black cohort in the underserved US Gulf South region of New Orleans, LA. Many subjects reported that enrollment in this study was the only healthcare contact they had related to post-COVID symptoms or concerns. The high prevalence of PASC symptoms reported is likely reflective of referral bias as multiple subjects reported that the current study was the only outlet they could find to discuss and document their symptoms. While this bias skews the reported prevalence data, the data is useful as an indicator of *which specific PASC symptoms are being reported* in this cohort and correlate clinical laboratory results. Fatigue and dyspnea were among the most reported symptoms, but problems with sleeping were reported to be the most frequent and severe PASC symptoms. PASC symptoms including fatigue and concentration issues were reported up to two years after symptom onset, underscoring the finding that PASC is not a brief or transient phenomenon for many participants. Clinical lab values were similar between PASC and non-PASC subjects, indicating an ongoing need for diagnostic biomarkers relevant across diverse patient groups. We found mild trends toward lower Hgb, Hct, and calcium in PASC vs non-PASC subjects, but population differences in Hgb and Hct were not observed when sex-specific cutoffs were used for these values. A limitation of the study is lack of pre-COVID data for these lab values, limiting the ability to attribute low values to PASC versus comorbidities.

Increased concentration of ECLIA numerical COI values of anti-SARS CoV-2 anti-N antibody over time after onset are reported in Roche ECLIA anti-N packet insert and, in the literature, [PMID: 34477548 and PMID 33708750], suggesting that COI value increase over time after onset can be used to reflect the immunity level of individuals after infection. For qualitative anti-N serum antibody using two unique assays, we did not find significant differences between non-PASC and PASC groups, though *among positive responders* we noted a trend toward higher quantitative anti-N response among PASC (vs non-PASC), subjects in the 61-120 days’ time bracket using the Roche assay. This may suggest a higher prevalence of re-infection in the PASC group as proposed by others [20] or support the hypothesis that long-lasting inflammation is due to continued presence of virus or viral particles in organs such as the gut [21] and consistent with the finding of higher IL-6 and TNF alpha by Olink, also reported as main biomarkers of PASC patients by others [22].

In summary, this 52.5% Black post-COVID cohort reports a high number of new-since-COVID symptoms, with similar symptoms reported across subjects, and a lack of clinical lab biomarker correlate in the lab panel tested. Initial research lab testing identifies a weak trend toward lower anti-N response in subjects with PASC (vs. non-PASC) and warrants further exploration considering emerging literature that PASC subjects may fail to eradicate a reservoir of virus, possibly in the gut. Future steps for this study will aim to further explore research laboratory results as currently available clinical laboratory testing does not appear to correlate with PASC and our cohort remains substantially impaired from PASC.

## Supporting information

Manuscript tables and supplemental figures

## Data Availability

All data produced in the present study are available upon reasonable request to the authors

## 5 Acknowledgements

This study was funded by Gilead IN-US-983-6062 and completed alongside the CDC-funded Clin-Seq-Ser study at University Medical Center and Tulane Medical Centers in New Orleans, LA.

## 6 Scope Statement

This work fits within the scope of infectious disease (ID) pathogenesis related to the emerging problem of post-acute sequelae of COVID-19 (PASC), which is increasingly recognized as a complication of COVID-19. The mechanism, diagnostic method, treatment, and long term prognosis for PASC remain unclear, warranting attention to this topic in the ID literature. The attached study identifies new-since-COVID symptoms in an underserved, highly comorbid, 53% Black population. The principal purpose of this study was to determine whether any result from a panel of commonly ordered clinical labs correlated with PASC in our cohort. In addition, we assessed whether acute COVID nasal/saliva SARS CoV-2 qRT-PCR or convalescent serum anti-SARS CoV-2 nucleocapsid correlated with PASC. Finally, we examined post-COVID nasal and saliva samples to determine whether there was evidence of post-COVID shedding in these compartments. We found that commonly ordered clinical labs do not correlate with PASC. We found no evidence that acute COVID respiratory PCR predicts PASC, nor evidence of prolonged respiratory shedding, but, among subjects with positive anti-N post COVID, a mild trend toward higher anti-N among PASC vs non-PASC, potentially supporting further evaluation with this assay in light of emerging concern for a cryptic intestinal reservoir.

## 7 Nomenclature

n.a.

## 9 Author Contributions

ES Data collection, data analysis, manuscript preparation, laboratory analysis AS Data analysis, manuscript preparation, laboratory analysis MM Data collection, data analysis, laboratory analysis, manuscript preparation MR Data collection, data analysis, laboratory analysis, manuscript preparation SI Data collection, data analysis, laboratory analysis, manuscript preparation UP Data collection, data analysis, laboratory analysis, manuscript preparation BC Data analysis, laboratory analysis, manuscript preparation SF Data collection, data analysis, laboratory analysis, manuscript preparation SL Data collection, data analysis, laboratory analysis, manuscript preparation MT Data collection, data analysis, laboratory analysis, manuscript preparation AL Data collection, data analysis, laboratory analysis, manuscript preparation CN Data collection, data analysis, laboratory analysis, manuscript preparation JM Data collection, data analysis, laboratory analysis, manuscript preparation JK Data collection, data analysis, laboratory analysis, manuscript preparation AD Study design and oversight, data collection, data analysis, manuscript preparation. DNF Study design and oversign, data collection data analysis, manuscript preparation.

## 11 Data Availability Statement

The datasets generated for this study can be found in the Supplemental data section. Additional data is available through direct request to the authors.

