## Supplementary material for "POST-ACUTE SEQUELAE OF COVID-19: CHARACTERIZATION, COMORBIDITIES, AND BIOMARKERS IN A DIVERSE COHORT": Manuscript tables and supplemental figures

Table 1. Cohort Characterization. Females in this cohort were 4.8 times as likely to report PASC positivity when compared to males. No significant odds ratios (ORs) were found when age, race, and BMI were stratified by sex

| Variable |  | PASC + N(%) | PASC - N(%) | OR (95% CI) | Pearson Chi-Square (p) |
| --- | --- | --- | --- | --- | --- |
| Age | 18-64 | 64 | 23 | 0.56 (0.11-2.73) | 0.533 (p=0.465) |
|  | 65+ | 10 | 2 |  |  |
| Sex | Female | 54 | 9 | 4.8 (1.83-12.59) | 11.039 (p < 0.001) |
|  | Male | 20 | 16 |  |  |
| Race | Black | 37 | 15 | 0.67 (0.27-1.67) | 0.749 (p= 0.387) |
|  | All other | 37 | 10 |  |  |
| BMI | <30 | 34 | 13 | 0.85 (0.34-2.11) | 1.173 (p=0.556) |
|  | ≥ 30 | 37 | 12 |  |  |

**PARTICIPANT**

Table 2. Chi square analysis of comorbidities/past medical history (PMH) and positivity for post acute sequelae of COVID-19 (PASC).

| **Pre-COVID Comorbidity** | **Pearson’s Chi-square value** | **P value** |
| --- | --- | --- |
| Cardiovascular | .225 | .635 |
| Hypertension | 2.443 | .118 |
| Hyperlipidemia | .081 | .777 |
| Diabetes | .042 | .837 |
| Pulmonary | .744 | .388 |
| Neurologic | .008 | .929 |
| Renal | 2.189 | .139 |
| Gastrointestinal | .132 | .717 |
| Liver | .346 | .556 |
| HIV | .100 | .752 |
| Heme (noncancer) | .037 | .847 |
| Cancer | .100 | .752 |
| Thyroid | .020 | .887 |
| Autoimmune | .776 | .378 |
| Hysterectomy | 1.382 | .240 |
| Tubal ligation or vasectomy | .350 | .554 |
| C section | 2.504 | .114 |
| Appendectomy | .699 | .403 |
| Cholecystectomy | .570 | .450 |
| Psychiatric | .060 | .807 |
| Orthopedic/pain condition | 1.024 | .312 |

Table 3. Severity of acute COVID-19 of post-COVID study subjects.

| **Severity Score** | **Clinical Course** | **N Participants with severity Score** |
| --- | --- | --- |
| 1 | Death | 0 |
| 2 | Hospitalized, on invasive mechanical ventilation or ECMO | 4 |
| 3 | Hospitalized, on non-invasive ventilation or high-flow O2 devices | 13 |
| 4 | Hospitalized, requiring supplemental O2 <15L | 22 |
| 5 | Hospitalized, not requiring supplemental O2 but requiring ongoing medical care | 8 |
| 6 | Hospitalized, not requiring O2, no longer requiring ongoing medical care | 1 |
| 7 | Not hospitalized | 23 |

Table 4a: Pearson’s Chi-square analysis of Maximum Severity Scores

|  | Max Severity score | | | | | |
| --- | --- | --- | --- | --- | --- | --- |
|  | 2 | 3 | 4 | 5 | 6 | 7 |
| PASC + | 4 | 12 | 11 | 5 | 1 | 38 |
| PASC - | 0 | 3 | 13 | 3 | 0 | 6 |
| Tot | 4 | 15 | 24 | 8 | 1 | 44 |
|  | Pearson Chi-square = 15.963, p= 0.007 | | | |  |  |

Table 4b: Independent samples T-test of Maximum Severity Scores

|  | N | Mean Severity Score | Std. Deviation | Std. Error Mean |
| --- | --- | --- | --- | --- |
| PASC + | 71 | 5.4225 | 1.83351 | 0.2176 |
| PASC - | 25 | 4.72 | 1.4 | 0.28 |

Table 5. Laboratory Value Descriptive Statistics.

| **Laboratory Value Descriptive Statistics** | | | | | | **Independent Samples t-test** | | |
| --- | --- | --- | --- | --- | --- | --- | --- | --- |
| Laboratory measure | PASC Positive? | N | Mean | Std. Deviation | Std. Error Mean | Equal variances assumed? | t-value | Two-sided p value |
| WBC | YES | 67 | 7.17 | 2.2 | 0.27 | Yes | -0.17 | 0.865 |
|  | NO | 24 | 7.27 | 2.8 | 0.58 |  |  |  |
| Hgb | YES | 67 | 13.51 | 1.4 | 0.17 | Yes | -2.601 | 0.011* |
|  | NO | 24 | 14.33 | 1.3 | 0.26 |  |  |  |
| HCT | YES | 67 | 41.49 | 3.7 | 0.45 | Yes | -2.843 | 0.006** |
|  | NO | 24 | 43.93 | 3.4 | 0.68 |  |  |  |
| Plt | YES | 67 | 279.01 | 71.0 | 8.68 | Yes | 1.227 | 0.223 |
|  | NO | 24 | 256.67 | 90.6 | 18.50 |  |  |  |
| Absolute neutrophils | YES | 67 | 4.21 | 1.8 | 0.23 | Yes | -0.014 | 0.989 |
|  | NO | 24 | 4.21 | 2.7 | 0.56 |  |  |  |
| % Neutrophils | YES | 67 | 57.15 | 10.1 | 1.23 | No | 0.853 | 0.4 |
|  | NO | 24 | 54.21 | 15.8 | 3.22 |  |  |  |
| Absolute lymphocyte | YES | 67 | 2.18 | 0.7 | 0.09 | Yes | -0.501 | 0.618 |
|  | NO | 24 | 2.27 | 0.8 | 0.16 |  |  |  |
| % Lymphocytes | YES | 67 | 31.42 | 8.8 | 1.07 | No | -1.011 | 0.32 |
|  | NO | 24 | 34.58 | 14.4 | 2.94 |  |  |  |
| ESR | YES | 67 | 16.49 | 14.8 | 1.80 | Yes | 0.573 | 0.568 |
|  | NO | 24 | 14.58 | 11.6 | 2.37 |  |  |  |
| Na | YES | 66 | 139.48 | 1.8 | 0.23 | No | 0.53 | 0.6 |
|  | NO | 23 | 139.13 | 3.0 | 0.63 |  |  |  |
| K | YES | 66 | 3.92 | 0.4 | 0.05 | Yes | -1.038 | 0.302 |
|  | NO | 23 | 4.01 | 0.4 | 0.09 |  |  |  |
| Cl | YES | 66 | 106.62 | 3.0 | 0.37 | Yes | 0.573 | 0.568 |
|  | NO | 23 | 106.17 | 3.8 | 0.80 |  |  |  |
| CO2 | YES | 66 | 27.79 | 2.8 | 0.34 | Yes | 0.585 | 0.56 |
|  | NO | 23 | 27.39 | 2.9 | 0.60 |  |  |  |
| Cr | YES | 66 | 0.91 | 0.3 | 0.04 | Yes | -1.699 | 0.093 |
|  | NO | 23 | 1.04 | 0.2 | 0.05 |  |  |  |
| Glucose | YES | 66 | 96.73 | 50.2 | 6.18 | No | -1.233 | 0.229 |
|  | NO | 23 | 122.22 | 94.6 | 19.73 |  |  |  |
| Calcium | YES | 66 | 9.25 | 0.4 | 0.05 | Yes | -2.307 | 0.023* |
|  | NO | 23 | 9.49 | 0.6 | 0.11 |  |  |  |
| HgbA1c | YES | 66 | 5.90 | 1.5 | 0.18 | Yes | -0.121 | 0.904 |
|  | NO | 24 | 5.94 | 1.5 | 0.31 |  |  |  |
| Ferritin | YES | 63 | 100.94 | 112.6 | 14.19 | Yes | -1.208 | 0.23 |
|  | NO | 23 | 134.59 | 119.0 | 24.81 |  |  |  |
| CPK | YES | 65 | 114.77 | 63.3 | 7.85 | No | -1.939 | 0.065 |
|  | NO | 23 | 199.83 | 207.0 | 43.15 |  |  |  |
| PT | YES | 63 | 10.97 | 1.6 | 0.21 | Yes | -0.067 | 0.947 |
|  | NO | 24 | 10.99 | 1.7 | 0.34 |  |  |  |
| PTT | YES | 65 | 27.94 | 3.2 | 0.40 | Yes | 1.577 | 0.118 |
|  | NO | 24 | 26.75 | 2.9 | 0.60 |  |  |  |
| Fibrinogen | YES | 66 | 372.26 | 111.1 | 13.68 | Yes | 0.521 | 0.604 |
|  | NO | 24 | 358.79 | 100.9 | 20.59 |  |  |  |
| CRP | YES | 66 | 0.64 | 0.8 | 0.10 |  |  |  |
|  | NO | 22 | 0.84 | 2.4 | 0.52 |  |  |  |
| High-Sensitivity Troponin | YES | 11 | 12.30 | 14.7 | 4.42 | Yes | 0.397 | 0.698 |
|  | NO | 4 | 9.25 | 4.6 | 2.29 |  |  |  |
| nt-Pro-BNP | YES | 64 | 174.00 | 584.0 | 73.00 | Yes | 0.805 | 0.423 |
|  | NO | 22 | 77.00 | 105.0 | 22.40 |  |  |  |
| INR | YES | 63 | 0.95 | 0.3 | 0.03 | Yes | 0.1 | 0.921 |
|  | NO | 24 | 0.95 | 0.3 | 0.06 |  |  |  |
| BUN | YES | 66 | 13.77 | 4.8 | 0.59 | Yes | -1.043 | 0.300 |
|  | NO | 23 | 15.00 | 5.0 | 1.04 |  |  |  |

**Supplemental**

Figure S1. SARS CoV-2 N1 Ct values from nasal swab qRT-PCR, individual subject results vs days from symptom onset.

SARS CoV-2 N1 Ct value (nasal)

| days from symptom onset to sample collection | CSS_005 |
| --- | --- |
| 7 | 34.1969 |
| 239 | 37.36 |
| 239 | 35.7 |
| 639 | 40 |
| 709 | 40 |

| days from symptom onset to sample collection | CSS_006 |
| --- | --- |
| 11 | 23.8986 |
| 663 | 40 |
| 663 | 40 |
| 754 | 40 |
| 754 | 40 |

| days from symptom onset to sample collection | CSS_016 |
| --- | --- |
| 19 | 29.102 |
| 19 | 29.744 |
| 243 | 40 |
| 243 | 36.3 |
| 705 | 40 |

| days from symptom onset to sample collection | CSS_022 |
| --- | --- |
| 20 | 31.92 |
| 20 | 32.739 |
| 647 | 40 |
| 738 | 40 |

| days from symptom onset to sample collection | CSS_024 |
| --- | --- |
| 666 | 40 |

| days from symptom onset to sample collection | CSS_033 |
| --- | --- |
| 10 | 40 |
| 10 | 17.986 |
| 14 | 24.702 |
| 14 | 24.705 |
| 713 | 40 |

| days from symptom onset to sample collection | CSS_041 |
| --- | --- |
| 216 | 40 |
| 650 | 40 |
| 650 | 40 |
| 727 | 40 |

| days from symptom onset to sample collection | CSS_055 |
| --- | --- |
| 37 | 36.672 |
| 37 | 40 |
| 717 | 40 |
|  | 17.564 |

| days from symptom onset to sample collection | CSS_095 |
| --- | --- |
| 579 | 35.9 |
| 579 | 40 |
| 670 | 40 |

| days from symptom onset to sample collection | CSS_097 |
| --- | --- |
| 636 | 40 |
| 723 | 40 |

| days from symptom onset to sample collection | CSS_121 |
| --- | --- |
| 97 | 40 |
| 665 | 40 |

| days from symptom onset to sample collection | CSS_225 |
| --- | --- |
| 9 | 40 |
| 9 | 31.4 |
| 9 | 40 |
| 382 | 40 |

| days from symptom onset to sample collection | CSS_229 |
| --- | --- |
| 5 | 34.016 |
| 5 | 35.9 |
| 382 | 40 |
|  | 40 |

| days from symptom onset to sample collection | CSS_231 |
| --- | --- |
| 445 | 40 |
| 579 | 40 |

| days from symptom onset to sample collection | CSS_253 |
| --- | --- |
| 550 | 40 |

| days from symptom onset to sample collection | CSS_254 |
| --- | --- |
| 689 | 32.1 |
| 689 | 40 |

| days from symptom onset to sample collection | CSS_258 |
| --- | --- |
| 8 | 33.296 |
| 445 | 40 |

| days from symptom onset to sample collection | CSS_263 |
| --- | --- |
| 10 | 40 |
| 444 | 40 |

| days from symptom onset to sample collection | CSS_272 |
| --- | --- |
| 5 | 26.2 |
| 219 | 36.961 |

| days from symptom onset to sample collection | CSS_295 |
| --- | --- |
| 8 | 34.284 |
| 8 | 40 |
| 356 | 40 |

Figure S2. SARS CoV-2 N1 Ct values from saliva qRT-PCR, individual subject results vs days from symptom onset.

SARS CoV-2 N1 Ct Value (saliva)

| days from symptom onset to sample collection | CSS_005 |
| --- | --- |
| 239 | 40 |
| 639 | 40 |
| 709 | 40 |

| days from symptom onset to sample collection | CSS_006 |
| --- | --- |
| 243 | 36.9 |
| 663 | 40 |
| 754 | 40 |

| days from symptom onset to sample collection | CSS_016 |
| --- | --- |
| 243 | 40 |
| 705 | 40 |

| days from symptom onset to sample collection | CSS_022 |
| --- | --- |
| 647 | 40 |
| 738 | 40 |

| days from symptom onset to sample collection | CSS_024 |
| --- | --- |
| 666 | 36.7 |

| days from symptom onset to sample collection | CSS_033 |
| --- | --- |
| 713 | 40 |

| days from symptom onset to sample collection | CSS_041 |
| --- | --- |
| 216 | 40 |
| 650 | 40 |
| 727 | 40 |

| days from symptom onset to sample collection | CSS_055 |
| --- | --- |
| 37 | 40 |
| 717 | 40 |

| days from symptom onset to sample collection | CSS_095 |
| --- | --- |
| 670 | 40 |

| days from symptom onset to sample collection | CSS_097 |
| --- | --- |
| 636 | 40 |
| 723 | 40 |

| days from symptom onset to sample collection | CSS_121 |
| --- | --- |
| 97 | 40 |
| 665 | 40 |

| days from symptom onset to sample collection | CSS_225 |
| --- | --- |
| 9 | 31.963 |
| 382 | 40 |

| days from symptom onset to sample collection | CSS_229 |
| --- | --- |
| 5 | 23.29 |
| 382 | 40 |
| 471 | 40 |

| days from symptom onset to sample collection | CSS_231 |
| --- | --- |
| 579 | 40 |

| days from symptom onset to sample collection | CSS_253 |
| --- | --- |
| 550 | 40 |

| days from symptom onset to sample collection | CSS_254 |
| --- | --- |
| 689 | 32.5 |

| days from symptom onset to sample collection | CSS_258 |
| --- | --- |
| 8 | 32.4 |
| 445 | 40 |

| days from symptom onset to sample collection | CSS_263 |
| --- | --- |
| 10 | 40 |
| 444 | 40 |

| days from symptom onset to sample collection | CSS_272 |
| --- | --- |
| 5 | 31.5 |

| days from symptom onset to sample collection | CSS_295 |
| --- | --- |
| 356 | 40 |

**Figure S3. Acute COVID-19 SARS CoV-2 N1 qRT-PCR results for individual subjects, grouped by later PASC status.** Nasal swab (a) and saliva (b) samples, grouped by later PASC status (black: no PASC, red: PASC), vs time between symptom onset and sample collection, presented for subjects with sample collected within 28d of symptom onset.

a.

SARS CoV-2 N1 PCR from acute nasal swabs, among later PASC vs non-PASC subjects, individual data

SARS CoV-2 N1 nasal Ct value

| days from symptom onset to sample | no PASC |
| --- | --- |
| 1 | 21.2 |
| 6 | 40 |
| 7 | 35.7 |
| 7 | 33.7 |
| 9 | 29.203 |
| 10 | 34.631 |
| 11 | 23.8986 |
| 15 | 27.942 |
| 15 | 27.53 |

| days from symptom onset to sample | PASC |
| --- | --- |
| 1 | 28.223 |
| 3 | 36.9 |
| 3 | 40 |
| 4 | 37.889 |
| 4 | 40 |
| 5 | 34.016 |
| 5 | 26.2 |
| 5 | 29.888 |
| 6 | 18.726 |
| 7 | 34.1969 |
| 7 | 40 |
| 7 | 30.782 |
| 7 | 37.825 |
| 8 | 33.296 |
| 8 | 34.284 |
| 9 | 31.4 |
| 9 | 40 |
| 9 | 40 |
| 10 | 17.986 |
| 10 | 40 |
| 10 | 32.1 |
| 10 | 33.3 |
| 11 | 35.916 |
| 11 | 36.157 |
| 12 | 31.3 |
| 14 | 24.705 |
| 16 | 17.564 |
| 17 | 34.242 |
| 19 | 29.102 |
| 20 | 31.92 |
| 26 | 40 |

b.

SARS CoV-2 N1 PCR from acute saliva, among later PASC vs non-PASC subjects, individual data

SARS CoV-2 N1 Ct Value (saliva)

| days from symptom onset to sample | no PASC |
| --- | --- |
| 1 | 18.6 |
| 1 | 26 |
| 6 | 29.7 |
| 7 | 36.2 |
| 7 | 26.7 |
| 9 | 24.339 |
| 10 | 32.504 |
| 15 | 37.421 |
| 15 | 40 |

| days from symptom onset to sample | PASC |
| --- | --- |
| 1 | 40 |
| 1 | 29.311 |
| 1 | 29.111 |
| 3 | 28.7 |
| 3 | 36.137 |
| 4 | 20.791 |
| 4 | 28.901 |
| 4 | 40 |
| 5 | 23.29 |
| 5 | 31.5 |
| 5 | 32.058 |
| 6 | 23.723 |
| 6 | 25.8 |
| 7 | 35.5 |
| 7 | 30.034 |
| 7 | 36.203 |
| 8 | 32.4 |
| 9 | 31.963 |
| 9 | 33.686 |
| 10 | 40 |
| 10 | 26.5 |
| 10 | 27.5 |
| 11 | 36.522 |
| 11 | 40 |
| 12 | 28.7 |
| 17 | 27.452 |

Table S1. Sequencing results. N=nasal, S=saliva.

CSS	Sample Type	Variant (posted on GISAID)

6	N	Predate Greek Naming

33	N	Predate Greek Naming

55	N	Predate Greek Naming

229	S	Predate Greek Naming

272	N	Predate Greek Naming

302	S	Predate Greek Naming

466	S	VOC Alpha 202012/01 GRY (B.1.1.7+Q.x)

486	S	VOC Delta GK/478K.V1 (B.1.617.2+AY.x)

535	N	VOC Delta GK/478K.V1 (B.1.617.2+AY.x)

602	S	VOC Delta GK/478K.V1 (B.1.617.2+AY.x)

604	S	VOC Delta GK/478K.V1 (B.1.617.2+AY.x)

718	S	VOC Delta GK/478K.V1 (B.1.617.2+AY.x)

725	N	VOC Delta GK/478K.V1 (B.1.617.2+AY.x)

Figure S4. Acute COVID infection VOC for patients who then presented for post-COVID study visits. a.Dominant variant of concern (VOC) circulating during acute COVID infection, for subjects who later enrolled as post-COVID subjects. x = VOC, y = number of subjects. b. Percent of subjects who are PASC positive grouped by their presumed acute COVID infection strain, based on time of symptom onset and dominant VOC at that time. x = VOC, y = percent of post COVID subjects whose acute COVID occurred during that VOC circulation and were PASC positive at post COVID visit.

a.

number of subjects

|  | VOC |
| --- | --- |
| predelta | 64 |
| delta | 34 |
| delta or omicron | 5 |
| omicron | 2 |

b.

percent

|  | Percent PASC Positive |
| --- | --- |
| pre delta | 74.1935 |
| delta | 71.875 |
| delta or omicron | 100 |

Figure S5. Serum anti-SARS CoV-2 nucleocapsid IgG results for PASC vs no PASC subjects using two distinct assays (Roche and, Zalgen), plotted versus days from symptom onset to blood draw. a. Roche anti-N COI (y) vs days from symptom onset to serum collection (x) for individual subjects. b. average Roche anti-N COI values per time bracket from symptom onset, excluding negative COI values. c. Zalgen anti-N OD/ml vs days from symptom onset to serum collection (y) for individual subjects. d. average Zalgen anti-N OD/ml values per time bracket from symptom onset. PASC=red, non-PASC=black. Bars=SEM. p value calcuated using student’s t test, 2 tails, unequal variance.

a.

Roche anti-N COI

| days from symptom onset to serum collection | PASC |
| --- | --- |
| 29 | 140.1 |
| 32 | 123.9 |
| 36 | 49.9 |
| 37 | 3.6 |
| 39 | 88.35 |
| 40 | 68.27 |
| 71 | 142.8 |
| 74 | 181.7 |
| 77 | 7.35 |
| 86 | 153.1 |
| 87 | 200.1 |
| 91 | 164.4 |
| 97 | 17.26 |
| 97 | 169.9 |
| 99 | 206.3 |
| 100 | 16.81 |
| 107 | 242.8 |
| 108 | 111 |
| 111 | 146.3 |
| 114 | 213.8 |
| 122 | 10.23 |
| 126 | 218.9 |
| 139 | 147.3 |
| 146 | 35.53 |
| 147 | 54.12 |
| 161 | 31.56 |
| 278 | 209.3 |
| 285 | 0.066 |
| 291 | 0.072 |
| 292 | 107.2 |
| 303 | 40.96 |
| 315 | 0.345 |
| 322 | 0.232 |
| 325 | 1.42 |
| 337 | 26.74 |
| 353 | 269.5 |
| 356 | 20.5 |
| 357 | 0.079 |
| 382 | 3.7 |
| 382 | 2.5 |
| 398 | 11.73 |
| 399 | 58.89 |
| 459 | 12.64 |
| 473 | 92.92 |
| 488 | 9.68 |
| 511 | 25.08 |
| 555 | 169 |
| 560 | 179.7 |
| 563 | 0.061 |
| 568 | 97.22 |
| 579 | 2.26 |
| 583 | 0.087 |
| 589 | 30.44 |
| 589 | 3.89 |
| 591 | 0.07 |
| 594 | 26.35 |
| 600 | 4.35 |
| 600 | 33.68 |
| 603 | 46.7 |
| 603 | 0.069 |
| 603 | 3.48 |
| 609 | 0.256 |
| 614 | 142.8 |
| 626 | 58 |
| 653 | 0.076 |
| 661 | 131.6 |
| 663 | 77.2 |
| 698 | 1.11 |

| days from symptom onset to serum collection | non PASC |
| --- | --- |
| 62 | 27.42 |
| 83 | 127.2 |
| 92 | 166.8 |
| 96 | 151.7 |
| 105 | 168.9 |
| 107 | 62.02 |
| 112 | 203.4 |
| 124 | 208.6 |
| 164 | 106 |
| 200 | 0.329 |
| 205 | 2.5 |
| 213 | 157.4 |
| 265 | 243.6 |
| 256 | 82.65 |
| 349 | 3.19 |
| 356 | 73.83 |
| 565 | 34.73 |
| 574 | 27.24 |
| 576 | 4.56 |
| 612 | 6.04 |
| 650 | 52.73 |
| 663 | 176.4 |

b.

Roche anti-N avg COI

| days from symptom onset to serum collection | PASC | no PASC |
| --- | --- | --- |
| D 0 to 60 | 79.02 |  |
| D 61-120 | 140.973 | 93.8571 |
| D 121 and up | 64.8157 | 84.2479 |

c.

Zalgen anti-N concentration (OD/ml)

| days from symptom onset to serum collection | PASC |
| --- | --- |
| 555 | 11.04 |
| 614 | 3.46 |
| 560 | 13.03 |
| 568 | 1.65307 |
| 594 | 2.17 |
| 626 | 3.05101 |
| 292 | 8.48 |
| 382 | 0.253332 |
| 382 | 3.20147 |
| 488 | 3.3702 |
| 698 | 6.79941 |
| 459 | 0.33 |
| 563 | 1.15661 |
| 353 | 12.8246 |
| 285 | 0.00973556 |
| 278 | 10.9125 |
| 300 | 4.65673 |
| 589 | 7.21 |
| 337 | 4.05221 |
| 603 | 1.68 |
| 589 | 0.72 |
| 139 | 13.0977 |
| 111 | 14.71 |
| 600 | 0.407095 |
| 86 | 55.4 |
| 122 | 0.6 |
| 114 | 36.61 |
| 356 | 1.00301 |
| 579 | 0.2 |
| 303 | 3.03441 |
| 97 | 1.46273 |
| 108 | 11.62 |
| 91 | 55.98 |
| 87 | 35.4 |
| 126 | 30.7208 |
| 591 | 0.22 |
| 398 | 0.9 |
| 74 | 38.21 |
| 71 | 54.01 |
| 99 | 22.1506 |
| 97 | 120 |
| 161 | 2.3169 |
| 77 | 6.8301 |
| 291 | 0.101649 |
| 36 | 0 |
| 37 | 0.41 |
| 600 | 1.35 |
| 325 | 1.49 |
| 603 | 0.14 |
| 147 | 5.33397 |
| 603 | 0.17 |
| 100 | 1.53711 |
| 107 | 48.4 |
| 653 | 0.310419 |
| 315 | 1.17223 |
| 322 | 0.472996 |
| 609 | 1.72209 |
| 357 | 0.415831 |
| 399 | 3.30248 |
| 29 | 9.10454 |
| 40 | 32.4278 |
| 39 | 39.2601 |
| 661 | 5.92483 |
| 146 | 2.24986 |
| 583 | 0.471926 |
| 32 | 120 |
| 663 | 2.69004 |
| 473 | 1.69615 |
| 511 | 0.57 |

| days from symptom onset to serum collection | no PASC |
| --- | --- |
| 663 | 3.18195 |
| 650 | 11.5758 |
| 565 | 4.73597 |
| 574 | 2.16 |
| 356 | 4.00476 |
| 576 | 0.18 |
| 349 | 1.06851 |
| 265 | 21.4178 |
| 213 | 30.4 |
| 164 | 9.04 |
| 112 | 46.03 |
| 92 | 31.4 |
| 200 | 0.3 |
| 312 | 1.73 |
| 105 | 55.74 |
| 96 | 23.14 |
| 124 | 58.8217 |
| 107 | 8.99441 |
| 83 | 22.8414 |
| 62 | 3.05 |
| 256 | 3.01 |
| 612 | 0.505532 |

d.

Zalgen anti-N avg concentration (OD/ml)

| days from symptom onset to serum collection | PASC | no PASC |
| --- | --- | --- |
| D 0 to 60 | 33.5337 |  |
| D 61-120 | 35.88 | 27.3137 |
| D 121 up | 3.71725 | 10.1421 |

**Table S2. O-link Urine and Plasma**

| **O-link Data in PASC positive vs. PASC negative Participants** | | | | | | |
| --- | --- | --- | --- | --- | --- | --- |
|  | **Plasma O-link** | | | **Urine O-link** | | |
|  | **t** | **degrees**  **of freedom** | **Two-Sided p** | **t** | **df** | **Two-Sided p** |
| **IL8** | **0.895** | **15** | **0.385** | **-0.073** | **9** | **0.943** |
| **VEGFA** | **-0.298** | **15** | **0.77** | **0.017** | **9** | **0.987** |
| **CD8A** | **0.468** | **15** | **0.646** | **1.169** | **9** | **0.272** |
| **MCP-3** | **1.003** | **15** | **0.332** | **1.157** | **9** | **0.277** |
| **GDNF** | **0.336** | **15** | **0.742** | **1.611** | **9** | **0.142** |
| **CDCP1** | **0.727** | **15** | **0.478** | **0.349** | **9** | **0.735** |
| **CD244** | **0.047** | **15** | **0.963** | **0.258** | **9** | **0.802** |
| **IL7** | **-0.353** | **15** | **0.729** | **-1.432** | **9** | **0.186** |
| **OPG** | **0.533** | **15** | **0.602** | **-0.707** | **9** | **0.497** |
| **LAP TGF-beta-1** | **-1.11** | **15** | **0.284** | **-1.003** | **9** | **0.342** |
| **uPA** | **0.08** | **15** | **0.937** | **-0.067** | **9** | **0.948** |
| **IL6** | **2.387** | **14.168** | **0.031*** | **1.725** | **9** | **0.119** |
| **IL-17C** | **-0.077** | **15** | **0.939** | **1.206** | **9** | **0.258** |
| **MCP-1** | **0.607** | **15** | **0.553** | **0.723** | **9** | **0.488** |
| **IL-17A** | **-0.733** | **15** | **0.475** | **0.529** | **9** | **0.61** |
| **CXCL11** | **-0.512** | **15** | **0.616** | **-0.74** | **9** | **0.478** |
| **AXIN1** | **-1.159** | **15** | **0.264** | **-0.696** | **9** | **0.504** |
| **TRAIL** | **0.78** | **15** | **0.447** | **0.226** | **9** | **0.826** |
| **IL-20RA** | **0.391** | **15** | **0.701** | **2.288** | **9** | **0.048** |
| **CXCL9** | **-0.208** | **15** | **0.838** | **0.687** | **9** | **0.509** |
| **CST5** | **-0.411** | **15** | **0.687** | **0.87** | **9** | **0.407** |
| **IL-2RB** | **0.363** | **15** | **0.722** | **0.827** | **9** | **0.429** |
| **IL-1 alpha** | **0.457** | **15** | **0.655** | **-1.313** | **9** | **0.222** |
| **OSM** | **-0.865** | **15** | **0.401** | **-0.31** | **9** | **0.764** |
| **IL2** | **0.828** | **15** | **0.421** | **1.721** | **9** | **0.119** |
| **CXCL1** | **-0.5** | **15** | **0.624** | **1.047** | **9** | **0.323** |
| **TSLP** | **-0.098** | **15** | **0.923** | **1.378** | **9** | **0.202** |
| **CCL4** | **0.762** | **15** | **0.458** | **0.849** | **9** | **0.418** |
| **CD6** | **-1.993** | **12.376** | **0.069** | **-0.503** | **9** | **0.627** |
| **SCF** | **1.133** | **15** | **0.275** | **1.512** | **9** | **0.165** |
| **IL18** | **0.929** | **15** | **0.368** | **-0.233** | **9** | **0.821** |
| **SLAMF1** | **0.161** | **15** | **0.874** | **0.88** | **4.675** | **0.422** |
| **TGF-alpha** | **-0.075** | **15** | **0.941** | **0.987** | **9** | **0.349** |
| **MCP-4** | **0.122** | **15** | **0.904** | **0.375** | **9** | **0.716** |
| **CCL11** | **-0.358** | **15** | **0.726** | **-0.163** | **9** | **0.874** |
| **TNFSF14** | **0.105** | **15** | **0.917** | **1.888** | **9** | **0.092** |
| **FGF-23** | **0.757** | **15** | **0.461** | **0.735** | **9** | **0.481** |
| **IL-10RA** | **0.913** | **15** | **0.376** | **-1.247** | **9** | **0.244** |
| **FGF-5** | **0.223** | **15** | **0.826** | **0.069** | **9** | **0.946** |
| **MMP-1** | **-0.638** | **15** | **0.533** | **0.905** | **9** | **0.389** |
| **LIF-R** | **1.284** | **15** | **0.219** | **-0.217** | **9** | **0.833** |
| **FGF-21** | **0.67** | **15** | **0.513** | **1.497** | **9** | **0.169** |
| **CCL19** | **1.474** | **15** | **0.161** | **-0.384** | **9** | **0.71** |
| **IL-15RA** | **0.289** | **15** | **0.776** | **2.022** | **9** | **0.074** |
| **IL-10RB** | **0.628** | **15** | **0.539** | **0.345** | **9** | **0.738** |
| **IL-22 RA1** | **1.421** | **15** | **0.176** | **1.606** | **4.806** | **0.171** |
| **IL-18R1** | **0.859** | **15** | **0.404** | **1.051** | **9** | **0.321** |
| **PD-L1** | **0.056** | **3.413** | **0.958** | **-0.233** | **9** | **0.821** |
| **Beta-NGF** | **-0.08** | **15** | **0.938** | **-2.99** | **9** | **0.015** |
| **CXCL5** | **-0.505** | **15** | **0.621** | **1.252** | **9** | **0.242** |
| **TRANCE** | **1.073** | **15** | **0.3** | **-1.565** | **9** | **0.152** |
| **HGF** | **0.405** | **15** | **0.691** | **-0.182** | **9** | **0.86** |
| **IL-12B** | **1.021** | **12.664** | **0.326** | **1.472** | **9** | **0.175** |
| **IL-24** | **-0.19** | **15** | **0.852** | **-0.371** | **9** | **0.72** |
| **IL13** | **0.78** | **15** | **0.447** | **1.626** | **9** | **0.138** |
| **ARTN** | **-0.808** | **3.034** | **0.478** | **0.788** | **9** | **0.451** |
| **MMP-10** | **-0.465** | **15** | **0.649** | **0.591** | **9** | **0.569** |
| **IL10** | **0.203** | **15** | **0.842** | **0.299** | **6.262** | **0.775** |
| **TNF** | **2.32** | **12.417** | **0.038*** | **-1.2** | **9** | **0.261** |
| **CCL23** | **0.492** | **15** | **0.63** | **0.164** | **9** | **0.873** |
| **CD5** | **0.059** | **15** | **0.953** | **-0.593** | **9** | **0.568** |
| **CCL3** | **0.903** | **15** | **0.381** | **0.454** | **9** | **0.661** |
| **Flt3L** | **0.537** | **15** | **0.599** | **0.821** | **9** | **0.433** |
| **CXCL6** | **0.523** | **15** | **0.608** | **0.237** | **9** | **0.818** |
| **CXCL10** | **1.006** | **15** | **0.33** | **-0.307** | **9** | **0.766** |
| **4E-BP1** | **-0.085** | **15** | **0.933** | **0.2** | **9** | **0.846** |
| **IL-20** | **0.445** | **15** | **0.662** | **-0.479** | **9** | **0.643** |
| **SIRT2** | **-0.206** | **15** | **0.839** | **1.653** | **9** | **0.133** |
| **CCL28** | **0.65** | **15** | **0.525** | **0.242** | **9** | **0.814** |
| **DNER** | **0.475** | **15** | **0.642** | **0.177** | **9** | **0.864** |
| **EN-RAGE** | **-0.738** | **15** | **0.472** | **-0.54** | **9** | **0.602** |
| **CD40** | **0.025** | **15** | **0.981** | **-0.262** | **9** | **0.799** |
| **IL33** | **0.723** | **15** | **0.481** | **0.411** | **9** | **0.691** |
| **IFN-gamma** | **1.143** | **15** | **0.271** | **1.306** | **9** | **0.224** |
| **FGF-19** | **-0.286** | **15** | **0.778** | **3.054** | **9** | **0.014** |
| **IL4** | **0.353** | **15** | **0.729** | **0.402** | **9** | **0.697** |
| **LIF** | **1.086** | **15** | **0.295** | **0.21** | **9** | **0.838** |
| **NRTN** | **1.033** | **15** | **0.318** | **-1.985** | **9** | **0.078** |
| **MCP-2** | **1.215** | **15** | **0.243** | **-0.396** | **9** | **0.701** |
| **CASP-8** | **0.682** | **15** | **0.506** | **0.68** | **9** | **0.514** |
| **CCL25** | **0.414** | **15** | **0.685** | **0.12** | **9** | **0.907** |
| **CX3CL1** | **0.858** | **15** | **0.404** | **0.363** | **9** | **0.725** |
| **TNFRSF9** | **0.918** | **15** | **0.373** | **-0.341** | **9** | **0.741** |
| **NT-3** | **1.025** | **15** | **0.322** | **2.21** | **7.414** | **0.061** |
| **TWEAK** | **0.504** | **15** | **0.622** | **-0.278** | **9** | **0.787** |
| **CCL20** | **1.567** | **15** | **0.138** | **0.718** | **9** | **0.491** |
| **ST1A1** | **-0.559** | **15** | **0.584** | **0.761** | **6.341** | **0.474** |
| **STAMBP** | **-0.5** | **15** | **0.625** | **-0.13** | **9** | **0.9** |
| **IL5** | **0.997** | **15** | **0.334** | **1.698** | **9** | **0.124** |
| **ADA** | **0.029** | **15** | **0.977** | **0.03** | **9** | **0.977** |
| **TNFB** | **0.941** | **15** | **0.362** | **0.886** | **9** | **0.399** |
| **CSF-1** | **1.546** | **15** | **0.143** | **0.101** | **9** | **0.922** |
